# Evolving Epidemiology of Stroke in India: Burden, Inequalities, and Risk Factors from 1990 to 2023 with Projections to 2035

**DOI:** 10.64898/2026.05.12.26352992

**Authors:** Manabesh Nath, Poorvi Tangri, Bhoomika Arora, Unnayni Joshi, Adila Jawaid, Kamalesh Kumar Patel, Ashish Datt Upadhyay, Awadh Kishor Pandit, Deepti Vibha, Pradeep Kumar

## Abstract

**Background and Purpose:** Stroke continues to be one of the major causes of death and long-term disability worldwide, with a greater impact in low- and middle-income countries. In India, there is limited evidence examining stroke burden and its changes over time and across regions. Therefore, we aimed to assess the burden of stroke in India from 1990 to 2023 using the latest data from the Global Burden of Disease (GBD) Study, along with projections up to 2035.

**Methods:** We used estimates from the GBD 2023 study to examine stroke incidence, prevalence, mortality, and disability-adjusted life years (DALYs) in India from 1990 to 2023. Age-standardized rates were analyzed to understand how these measures have changed over time. We also conducted state-level analyses to explore regional differences in stroke burden. The contributions of all major modifiable risk factors were assessed using population-attributable fractions. In addition, we projected future trends in stroke burden up to 2035.

**Results:** From 1990 to 2023, the percentage change in overall stroke burden in India showed minimal variation across key indicators. Incidence remained largely stable (0.00% [–0.04 to 0.05]), while prevalence showed a slight increase (0.06% [0.03 to 0.10]). Mortality (–0.11% [–0.36 to 0.20]) and DALYs (–0.17% [–0.38 to 0.12]) demonstrated modest declines over the study period. Notable regional disparities were evident, with states such as Chhattisgarh, Assam, and Jharkhand bearing the highest burden. High systolic blood pressure remained the leading risk factor in 2023, contributing the largest share of stroke-related deaths, followed by dietary risks, air pollution, tobacco use, and high body mass index. Future projections indicate that by 2035, stroke prevalence is likely to increase, while incidence, mortality, and DALYs are expected to show only modest changes.

**Conclusions:** Stroke remains a major and growing public health challenge in India with a continuing increase in burden despite slight improvements in age-standardized rates over time. Addressing this challenge will require stronger prevention efforts, better control of key risk factors, and focused strategies to reduce regional disparities in stroke burden nationwide.

## Introduction

Stroke continues to be one of the leading causes of death and long-term disability worldwide, despite significant advances in its prevention, diagnosis, and treatment.^1^ In recent decades, there has been a major shift in global health patterns, with non-communicable diseases becoming the primary drivers of illness and mortality.^2^ Among these, stroke stands out as a major public health concern due to its wide-ranging clinical, economic, and social impact on individuals and healthcare systems.^3^ According to the Global Burden of Disease (GBD) Study, stroke ranked as the second leading cause of death and the third leading cause of combined death and disability globally in 2021, contributing to millions of deaths and a substantial share of disability-adjusted life years (DALYs) worldwide.^4^

Although age-standardized stroke mortality rates have declined in many high-income countries—largely due to better control of risk factors, increased public awareness, and advances in acute stroke care—the overall global burden of stroke continues to rise.^5^ This increase is mainly driven by population growth, aging, and a growing prevalence of modifiable risk factors, particularly in low- and middle-income countries. Evidence from the GBD Study shows that the number of people living with cardiovascular diseases, including stroke, increased markedly from about 271 million in 1990 to 523 million in 2019, reflecting the expanding global burden.^6^ In addition, while age-standardized cardiovascular mortality declined during the late twentieth century, recent trends indicate that this progress has slowed in several regions, with some countries even experiencing stagnation or reversal of earlier gains.^7^

An increasing prevalence of both modifiable and non-modifiable risk factors largely drives the rising burden of stroke.^8^ Key contributors include hypertension, diabetes, overweight and obesity, unhealthy diets, tobacco use, and physical inactivity, along with environmental exposures such as air pollution and the effects of population aging.^9^ Evidence from the GBD Study indicates that nearly 90% of the global stroke burden can be attributed to a combination of behavioral, metabolic, and environmental risk factors.^10^ This highlights the importance of closely monitoring these factors to inform effective prevention strategies and public health policies. The World Health Organization (WHO) has emphasized the need to address these upstream determinants—particularly blood pressure control, diabetes management, tobacco cessation, and promotion of healthy lifestyles—as key priorities for reducing stroke risk and improving overall population health.^11^

The GBD Study provides a comprehensive, standardized framework for assessing the burden of disease and its risk factors across countries and over time.^12^ By integrating data from multiple sources—including disease surveillance systems, vital registration records, administrative health databases, and epidemiological studies—the GBD generates estimates of incidence, prevalence, mortality, and DALYs for a wide range of conditions.^13,14^ This standardized approach enables meaningful comparisons across regions and time periods. Recent GBD analyses have revealed significant geographic variation in stroke burden, driven by differences in socioeconomic development, access to healthcare, and exposure to risk factors.^1,5^ To better interpret these variations, regions are often categorized using the Sociodemographic Index (SDI), a composite measure that includes income per capita, educational attainment, and fertility rates, reflecting levels of development and stages of epidemiological transition.

India, which accounts for nearly one-sixth of the global population, is undergoing rapid demographic, epidemiological, and socioeconomic changes that are likely to influence the burden and distribution of stroke.^15,16^ Over the past three decades, the country has seen substantial population growth, increased life expectancy, rapid urbanization, and shifts in lifestyle behaviors, all of which have contributed to greater exposure to stroke risk factors.^17,18^ Although previous studies have reported rising trends in stroke incidence and mortality in India, comprehensive long-term analyses that examine temporal patterns, regional disparities, stroke subtypes, and attributable risk factors at both national and state levels remain limited.^19–22^ Given India’s vast geographic diversity and significant variations in healthcare access, socioeconomic conditions, environmental exposures, and lifestyle practices, understanding these evolving patterns across states is crucial for guiding effective health policies and resource allocation.^23–26^

Several studies from India have explored individual risk factors for stroke. Still, only a few have provided a comprehensive assessment of the state-wise burden and the contribution of major risk factors across different stroke subtypes, including ischaemic stroke, intracerebral haemorrhage, and subarachnoid haemorrhage.^27–35^ Stroke is largely preventable through modification of key risk factors such as hypertension, tobacco use, unhealthy diet, physical inactivity, and metabolic disorders.^36^ Therefore, understanding how stroke burden and its risk factors vary across regions is essential for developing targeted prevention strategies and implementing region-specific public health interventions.

In this context, the present study aims to provide a comprehensive assessment of the changing epidemiology of stroke in India from 1990 to 2023 using estimates from the Global Burden of Disease Study. We examine trends in incidence, prevalence, mortality, and DALYs across age groups, sex, geographic regions, and stroke subtypes. We also evaluate state-wise variations in stroke burden and quantify the contribution of major modifiable risk factors, including hypertension, tobacco use, metabolic factors, dietary risks, and environmental exposures. In addition, we project the future burden of stroke in India up to 2035. By generating updated national and subnational estimates, risk attribution, and projections, this study aims to support evidence-based policymaking and guide strategies to reduce the growing burden of stroke in India.

## Methods

### Study Design and Data Source

This study is a secondary analysis of data from the GBD Study 2023,^37^ which provides comprehensive and comparable estimates of disease burden across countries, regions, and over time. The GBD framework integrates data from multiple sources—including vital registration systems, population-based disease registries, administrative health databases, hospital records, national surveys, and published epidemiological studies—to generate standardized estimates of incidence, prevalence, mortality, and DALYs.

For this analysis, we extracted data for India from 1990 to 2023, including both national and state-level estimates where available. The study examined the burden of overall stroke as well as major stroke subtypes, including ischaemic stroke (IS), intracerebral hemorrhage (ICH), and subarachnoid hemorrhage (SAH). Estimates were analyzed across age groups (15–49 years, 50–69 years, and ≥70 years), geographic regions (states), and time periods (1990–2000, 2000–2010, 2010–2023, and the overall period 1990–2023).

### Case Definition and Stroke Subtypes

Stroke was defined according to WHO clinical criteria^38^ as the rapid development of clinical signs of focal or global disturbance of cerebral function lasting more than 24 hours or leading to death, with no apparent cause other than a vascular origin.²

Within the GBD framework, stroke is categorized into three main subtypes:

a. **Ischaemic stroke:** caused by obstruction of cerebral blood flow due to thrombotic or embolic events.
b. **Intracerebral hemorrhage:** resulting from bleeding within the brain tissue.
c. **Subarachnoid hemorrhage:** caused by bleeding into the subarachnoid space surrounding the brain.

### Burden Measures

The burden of stroke was assessed using key epidemiological indicators to capture both its frequency and overall health impact:

1. **Incidence:** the number of new stroke cases occurring within a specified time period.
2. **Prevalence:** the total number of individuals living with stroke at a given point in time.
3. **Mortality:** the number of deaths attributable to stroke.
4. **DALYs:** a summary measure that combines years of life lost due to premature death with years lived with disability, reflecting the overall disease burden.

Both **absolute numbers** and **age-standardized rates per 100,000 population** were analyzed. Age standardization was used to allow meaningful comparisons across time periods and geographic regions by accounting for differences in population age structure.

### Risk Factor Attribution

To estimate the contribution of modifiable risk factors to the stroke burden, we used population-attributable fractions (PAFs) derived from the GBD comparative risk assessment framework.³ This method quantifies the proportion of disease burden that can be attributed to specific risk factors by combining information on population exposure levels with relative risks obtained from epidemiological studies.

The analysis included a broad range of major modifiable risk factors^39^:

a. **High systolic blood pressure**
b. **Dietary risks**, including diets low in fruits, vegetables, whole grains, fiber, and omega-6 PUFA, and diets high in red meat, processed meat, sodium, and sugar-sweetened beverages
c. **Alcohol use, smoking, and overall tobacco use**
d. **High body mass index**
e. **High fasting plasma glucose**
f. **Air pollution**, including ambient particulate matter pollution and household air pollution from solid fuels
g. **Temperature-related exposures**, including high and low temperatures
h. **Physical inactivity**
i. **High low-density lipoprotein (LDL) cholesterol**

### Geographic and State-Level Analysis

To explore regional differences, we analyzed stroke burden across Indian states and union territories. State-level estimates were assessed for incidence, prevalence, mortality, DALYs, and the contribution of key risk factors. This approach enabled us to identify geographic variations in stroke burden and differences in exposure to major risk factors across regions.

### Statistical Analysis

Temporal trends in stroke burden from 1990 to 2023 were evaluated using age-standardized incidence, prevalence, mortality, and DALY rates. Percentage changes over time were calculated to understand the magnitude and direction of change in disease burden during the study period.

All estimates were reported with 95% uncertainty intervals (UIs), as provided by the GBD study, to account for variability arising from data sources and modeling methods. Data extraction and analysis were conducted using R statistical software (version 4.5.2; R Foundation for Statistical Computing, Vienna, Austria).^40^

### Projection of Stroke Burden to 2035

Future projections of stroke burden in India, including incidence, prevalence, mortality, and DALYs, were generated up to 2035 using linear regression models based on historical trends. The dataset was imported from Excel using the *readxl* package and analyzed in R. Separate models were fitted for each outcome, with calendar year as the independent variable, and their corresponding uncertainty bounds (value, upper, and lower estimates).

Predictions were generated for future time points (2025, 2030, and 2035) using the predict() function. These projected values were then combined with observed data using the *dplyr* package. Multi-panel visualizations were created using *ggplot2*, incorporating shaded uncertainty intervals and clearly distinguishing between observed and projected trends.^41^

## Results

### Overall Burden of Stroke in India

In 2023, stroke remained a major public health concern in India, accounting for approximately 1.87 million incident cases (95% CI: 1.67–2.10 million) and 13.64 million prevalent cases (95% CI: 12.38–15.03 million). It was responsible for an estimated 1.03 million deaths (95% CI: 0.83–1.26 million) and 25.99 million disability-adjusted life years (DALYs) (95% CI: 20.48–31.67 million). The age-standardized incidence rate was 151.66 per 100,000 population (95% CI: 135.96–170.55), while the age-standardized mortality rate was 92.88 per 100,000 population (95% CI: 74.42–112.97). Overall, stroke contributed substantially to the burden of non-communicable diseases in India, accounting for 10.51% of total deaths (95% CI: 8.58–12.52%) and 5.38% of total DALYs (95% CI: 4.30–6.58).

Across stroke subtypes, ischaemic stroke accounted for the largest proportion of cases, followed by intracerebral hemorrhage and subarachnoid hemorrhage. In 2023, the estimated number of cases was 1.09 million (95% CI: 0.94–1.28 million) for ischaemic stroke, 0.65 million (95% CI: 0.54–0.75 million) for intracerebral hemorrhage, and 0.13 million (95% CI: 0.11–0.14 million) for subarachnoid hemorrhage (**Table 1**).

**Table 1.**
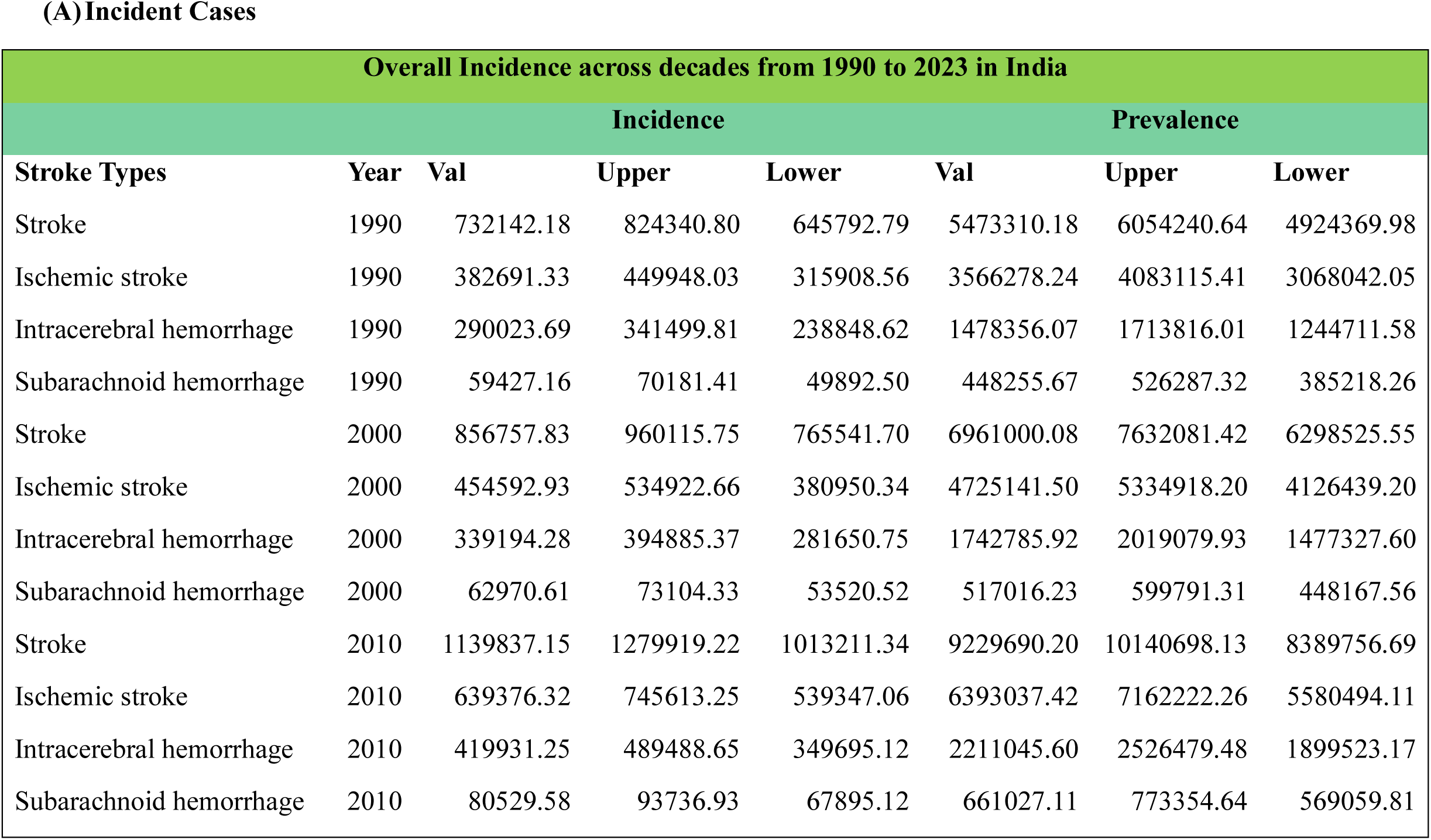
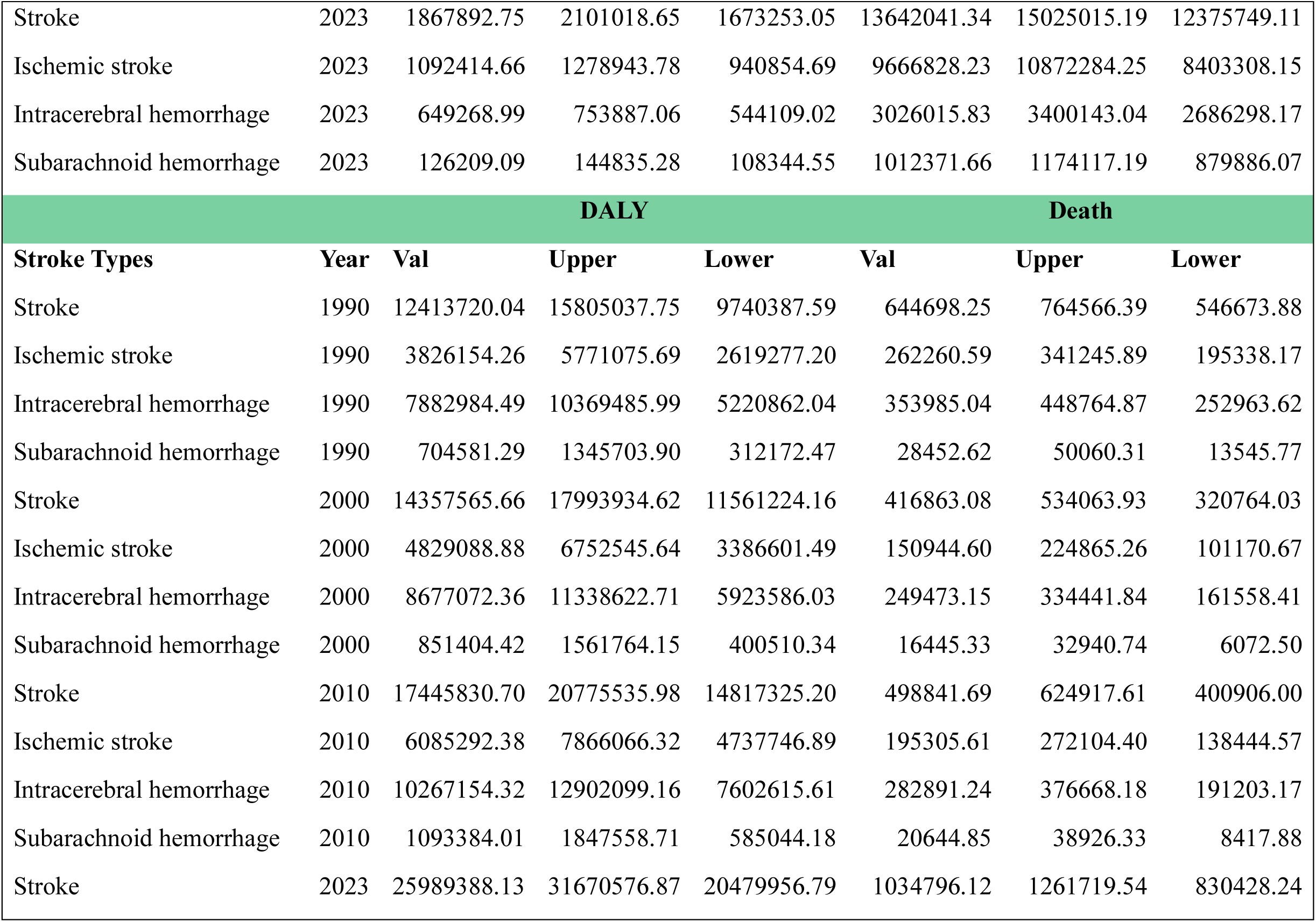
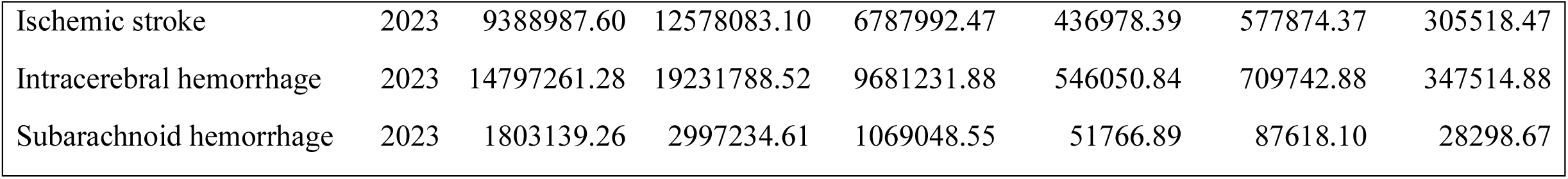
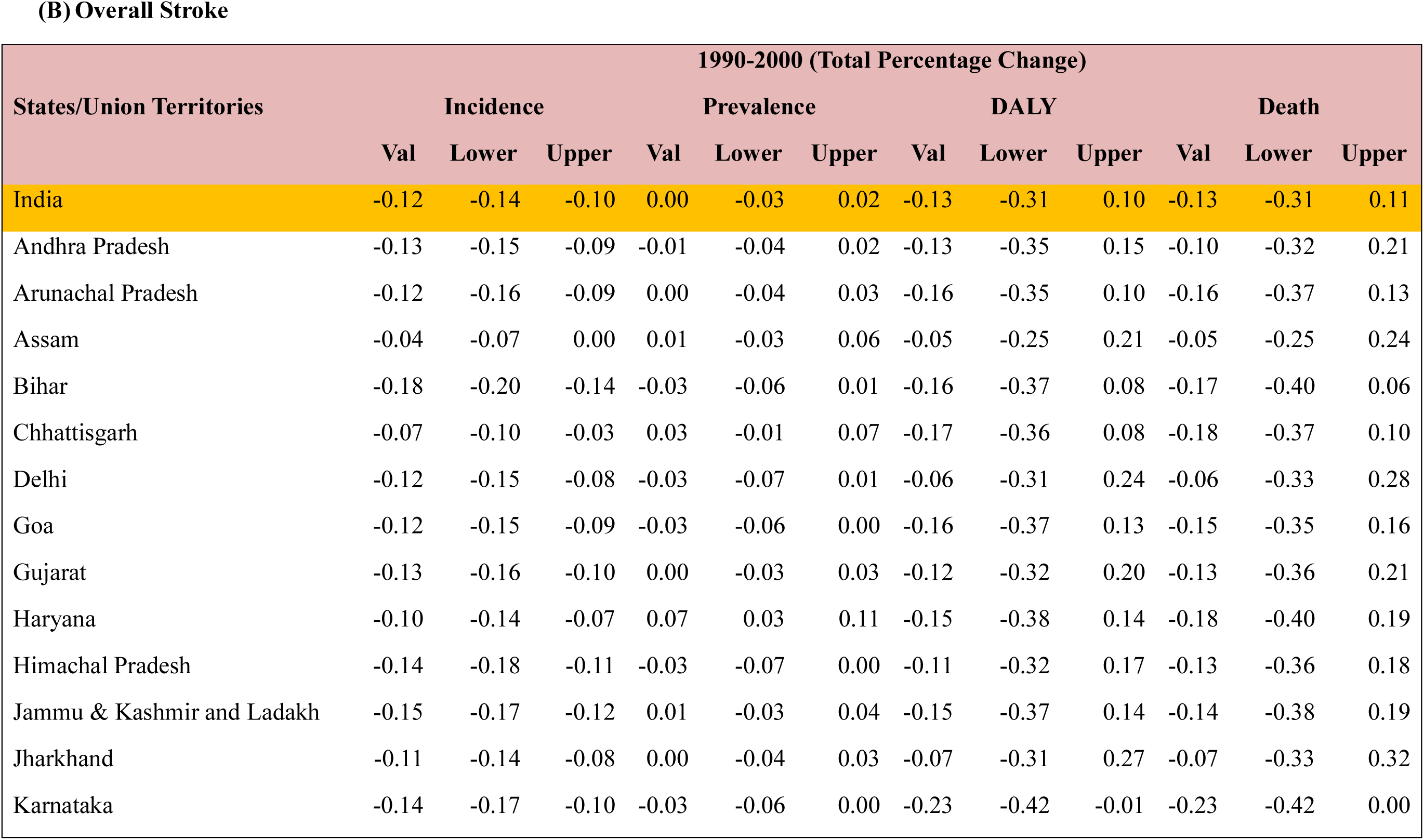
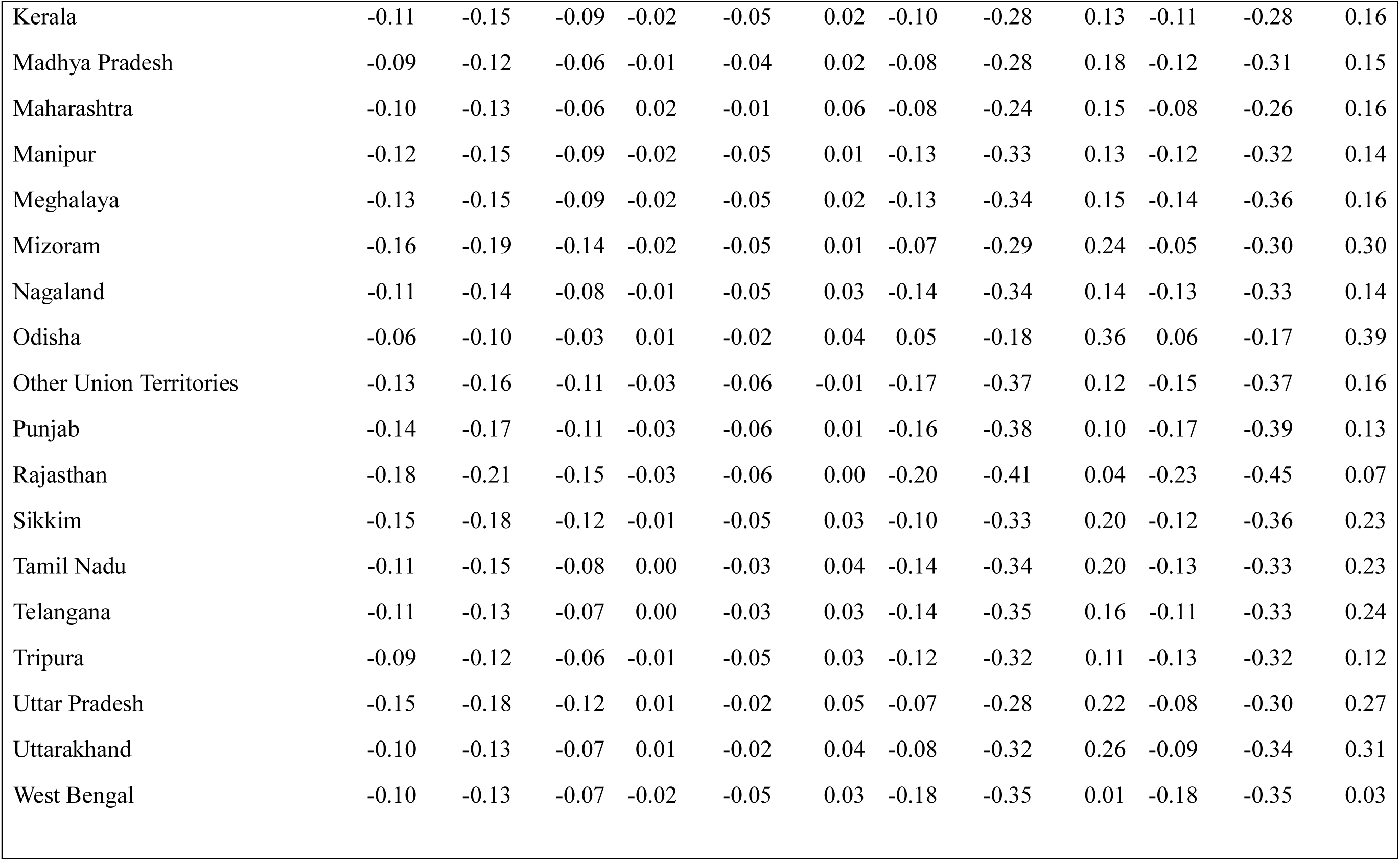
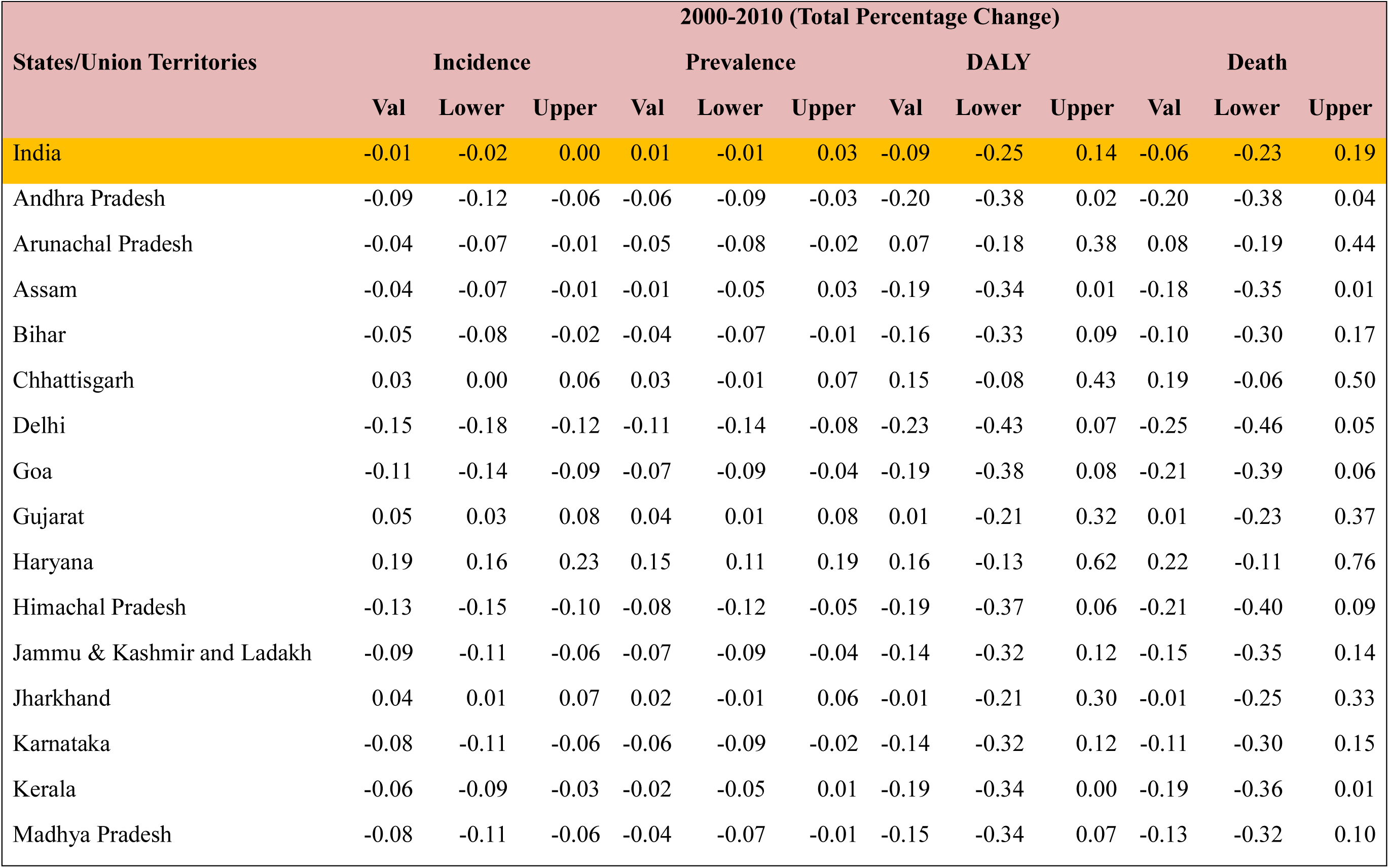
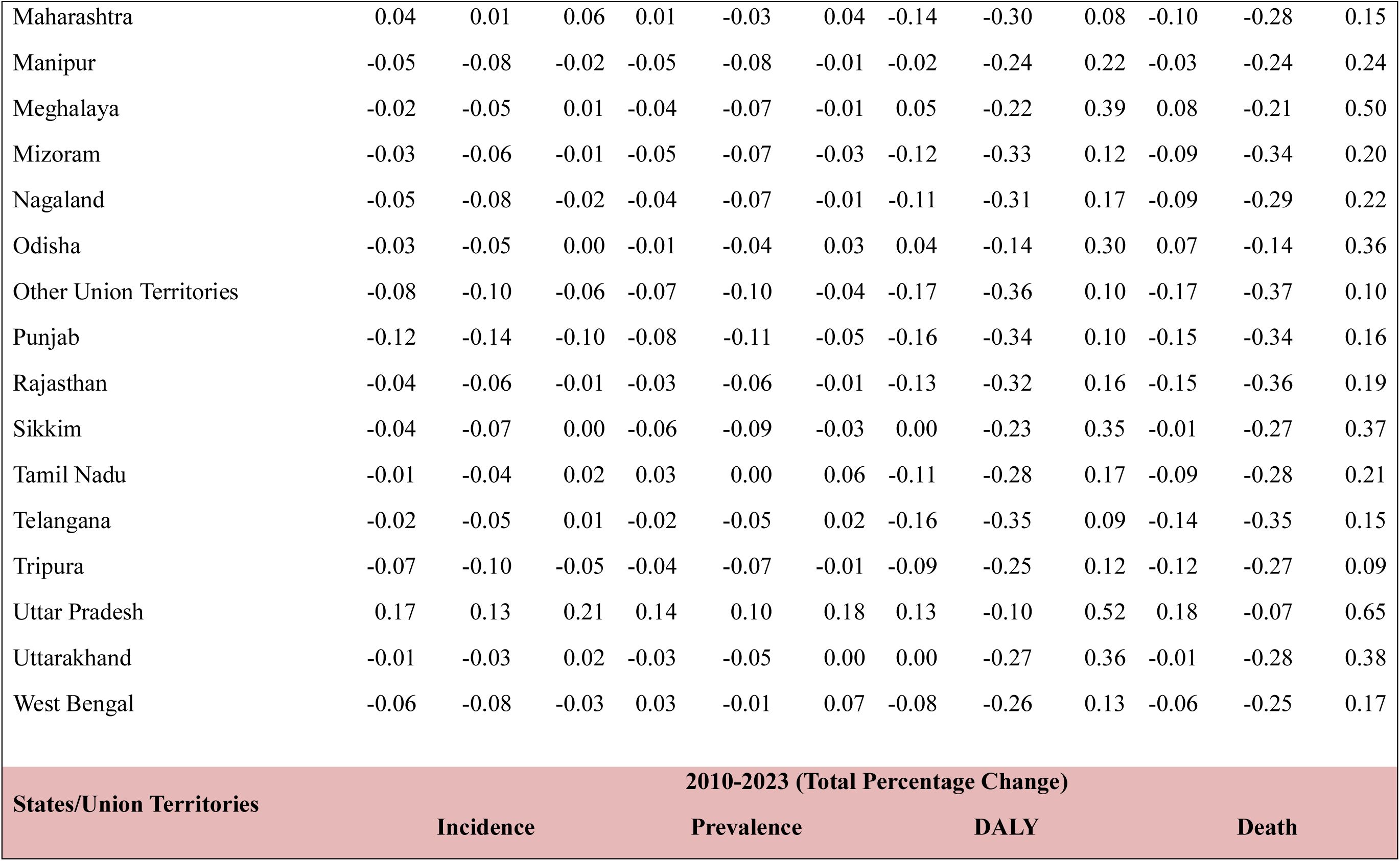
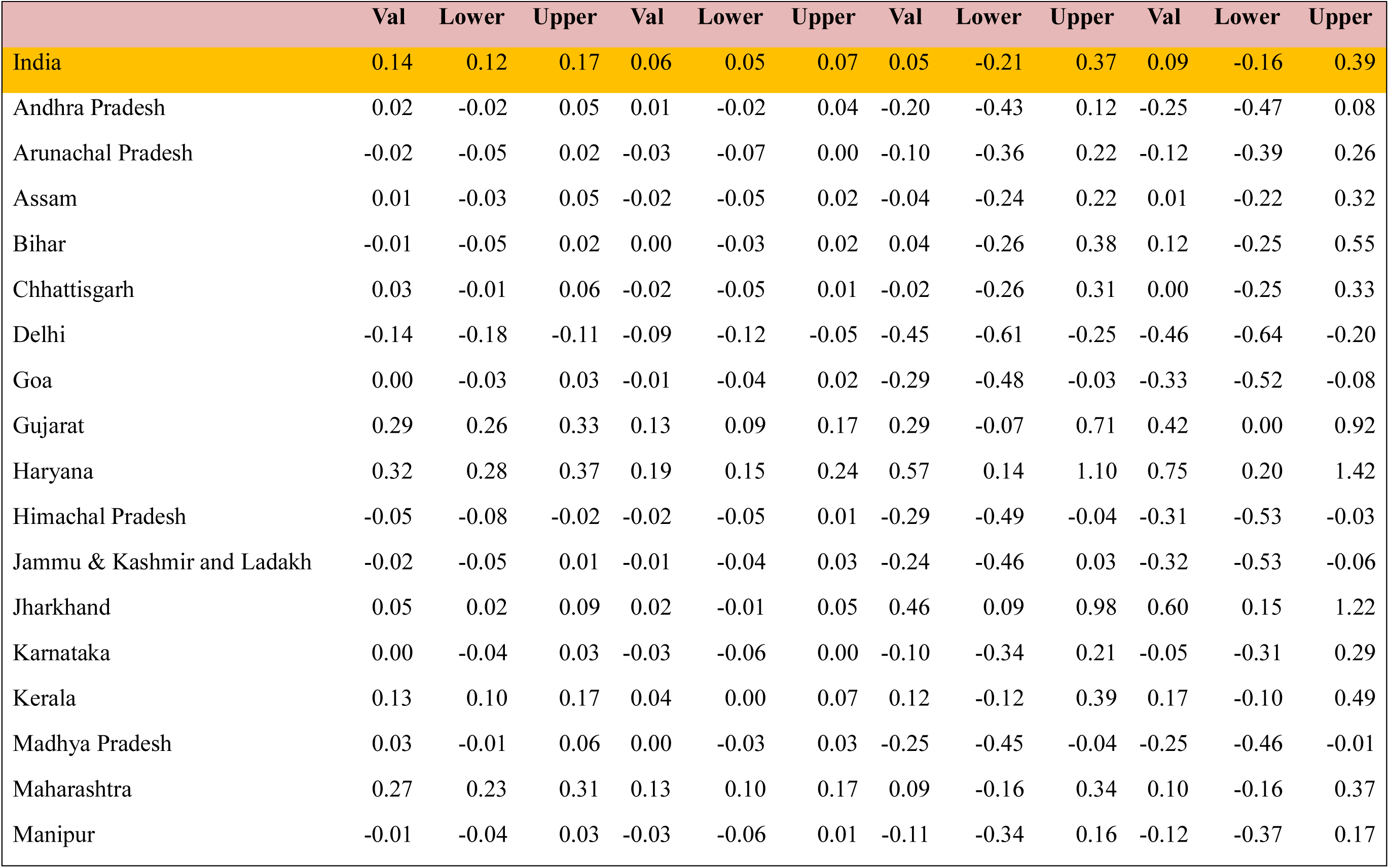
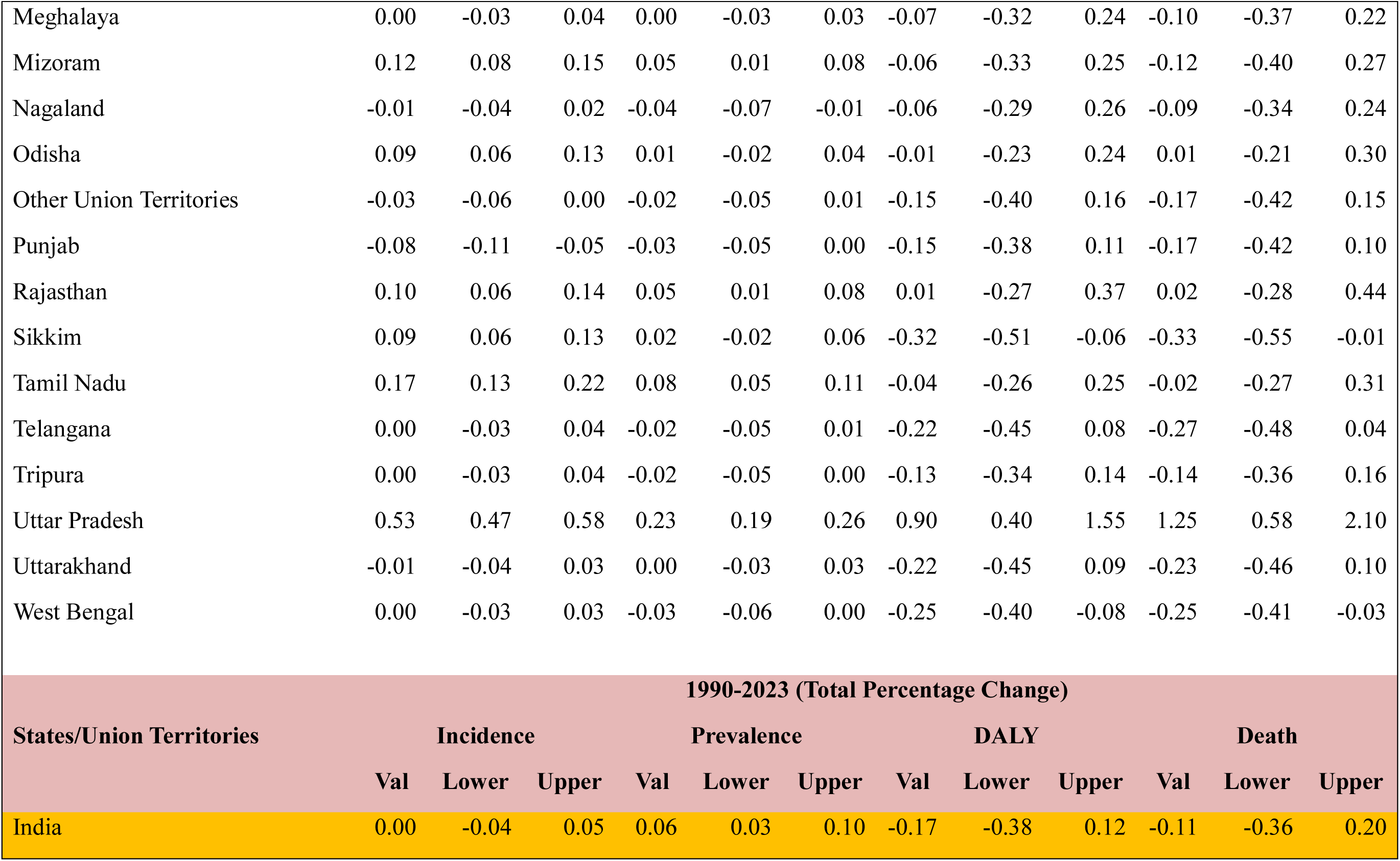
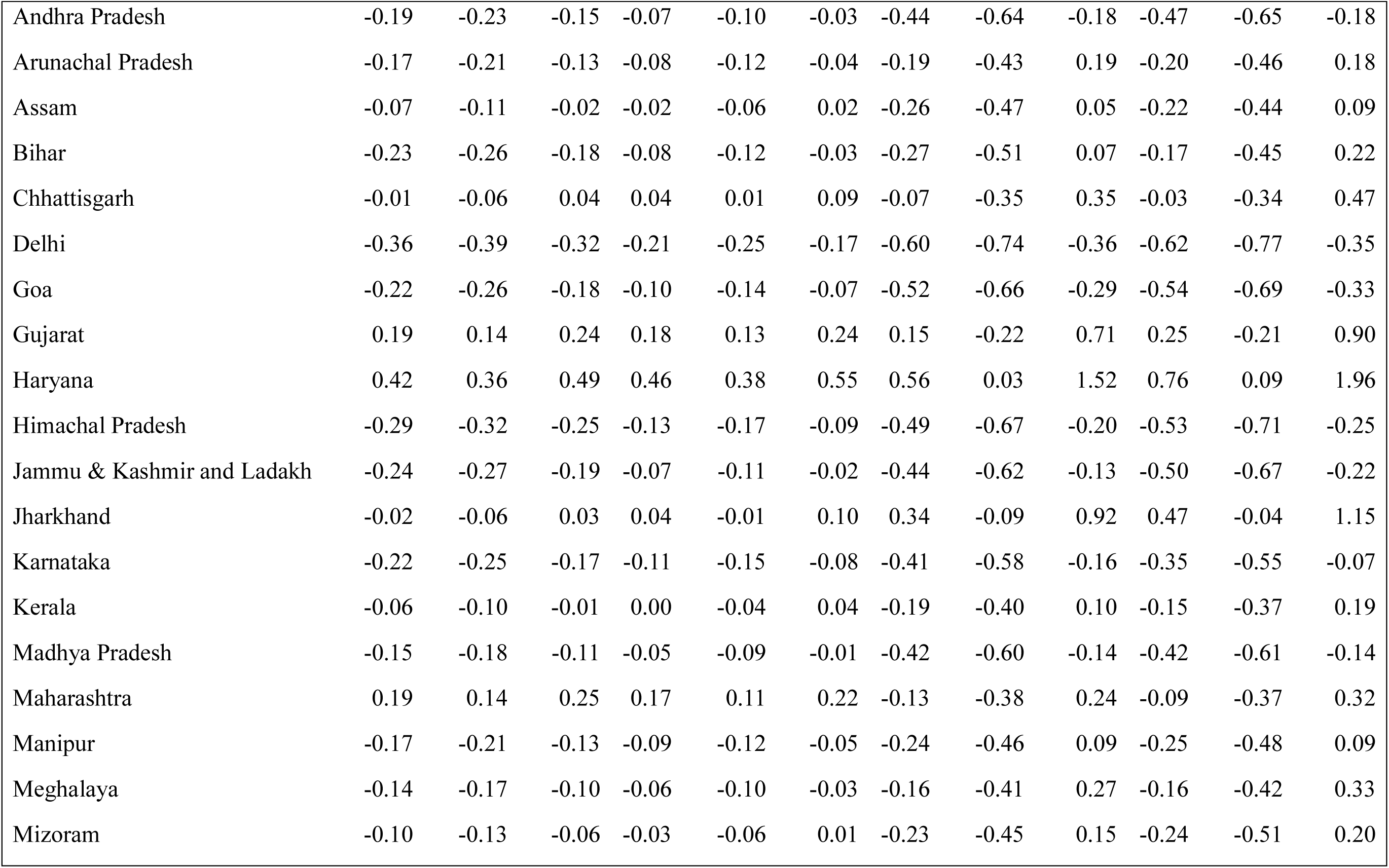
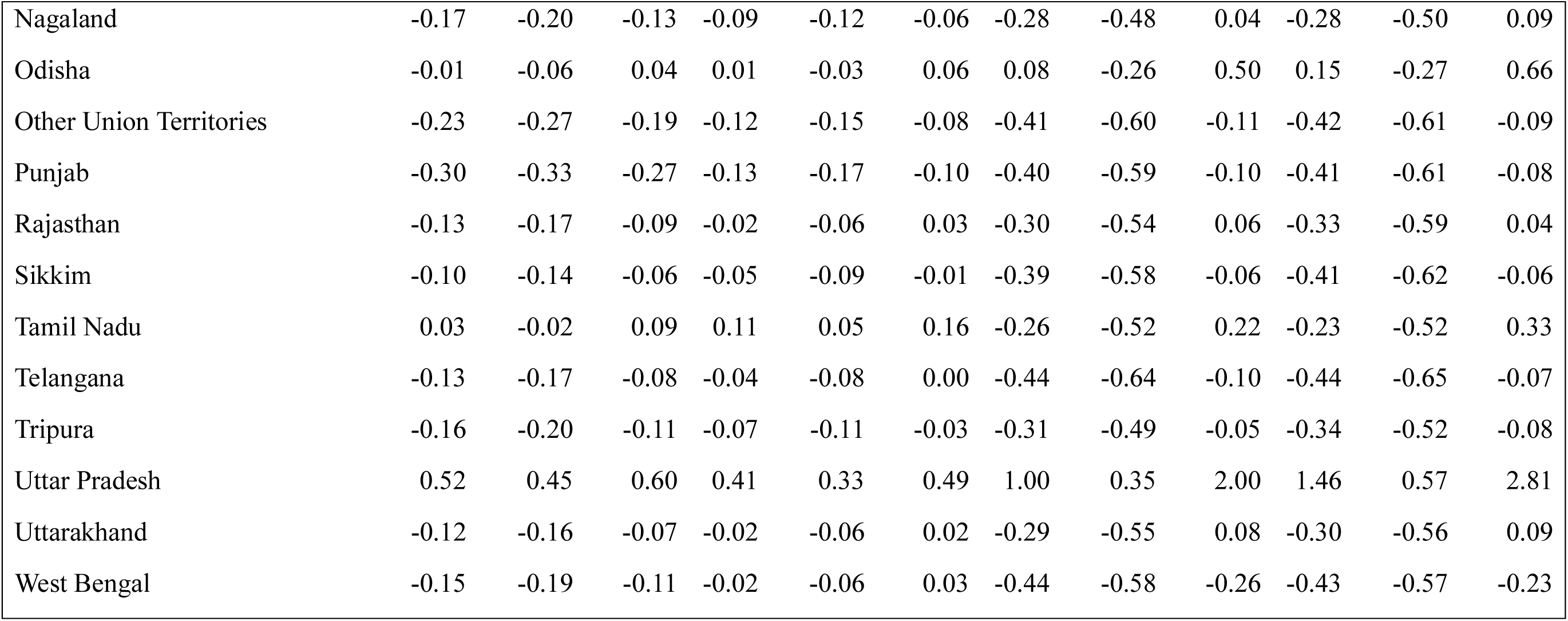
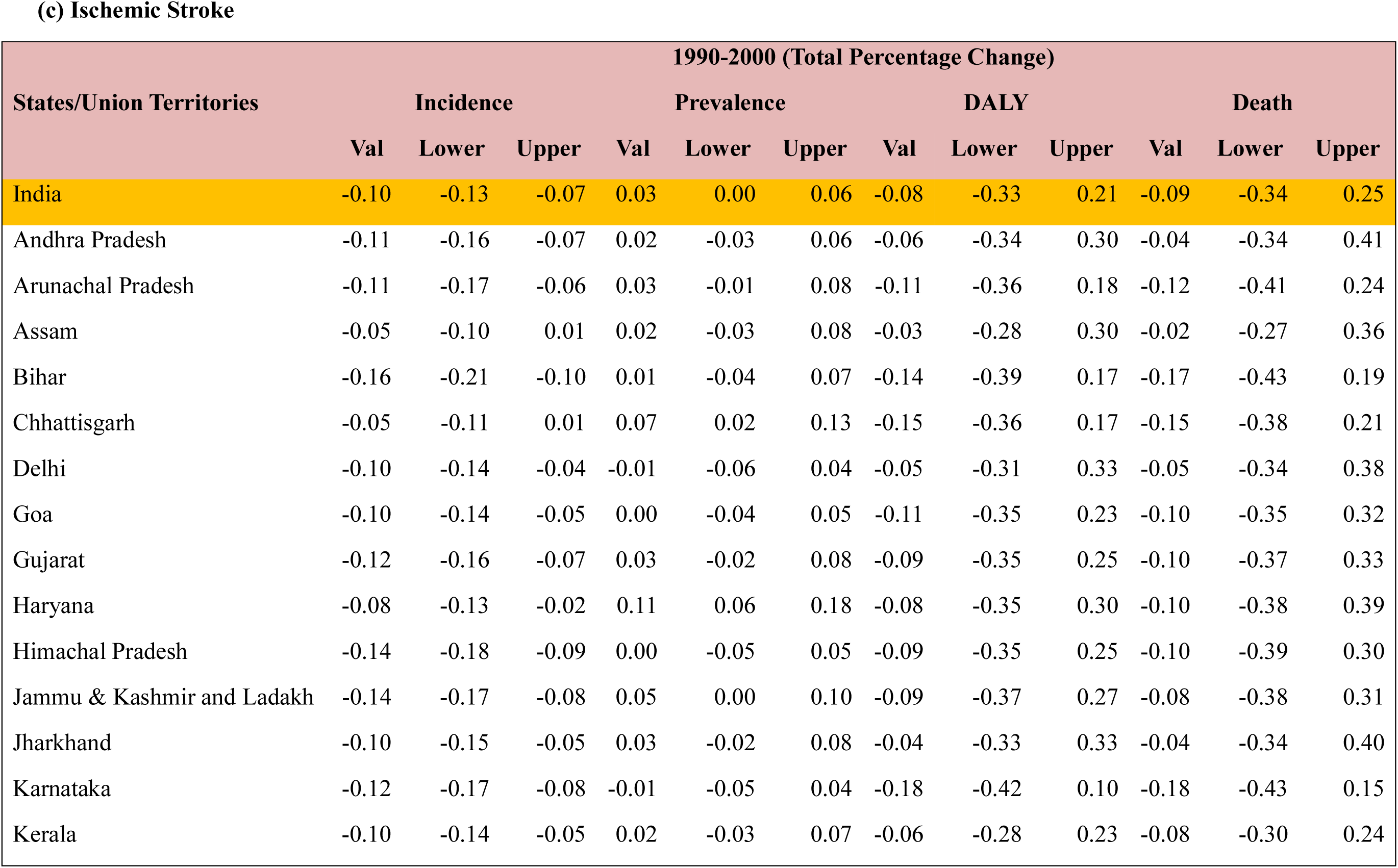
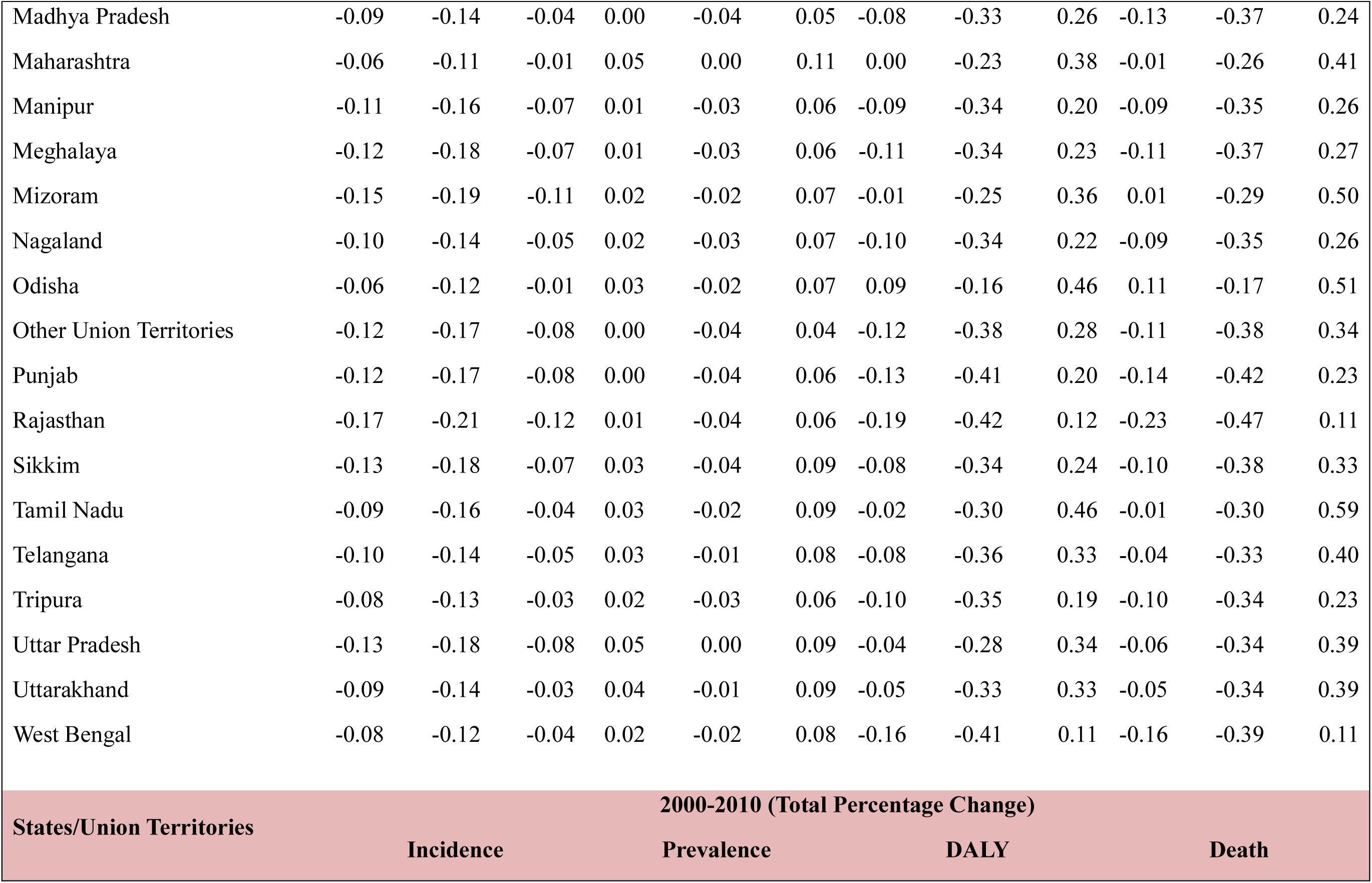
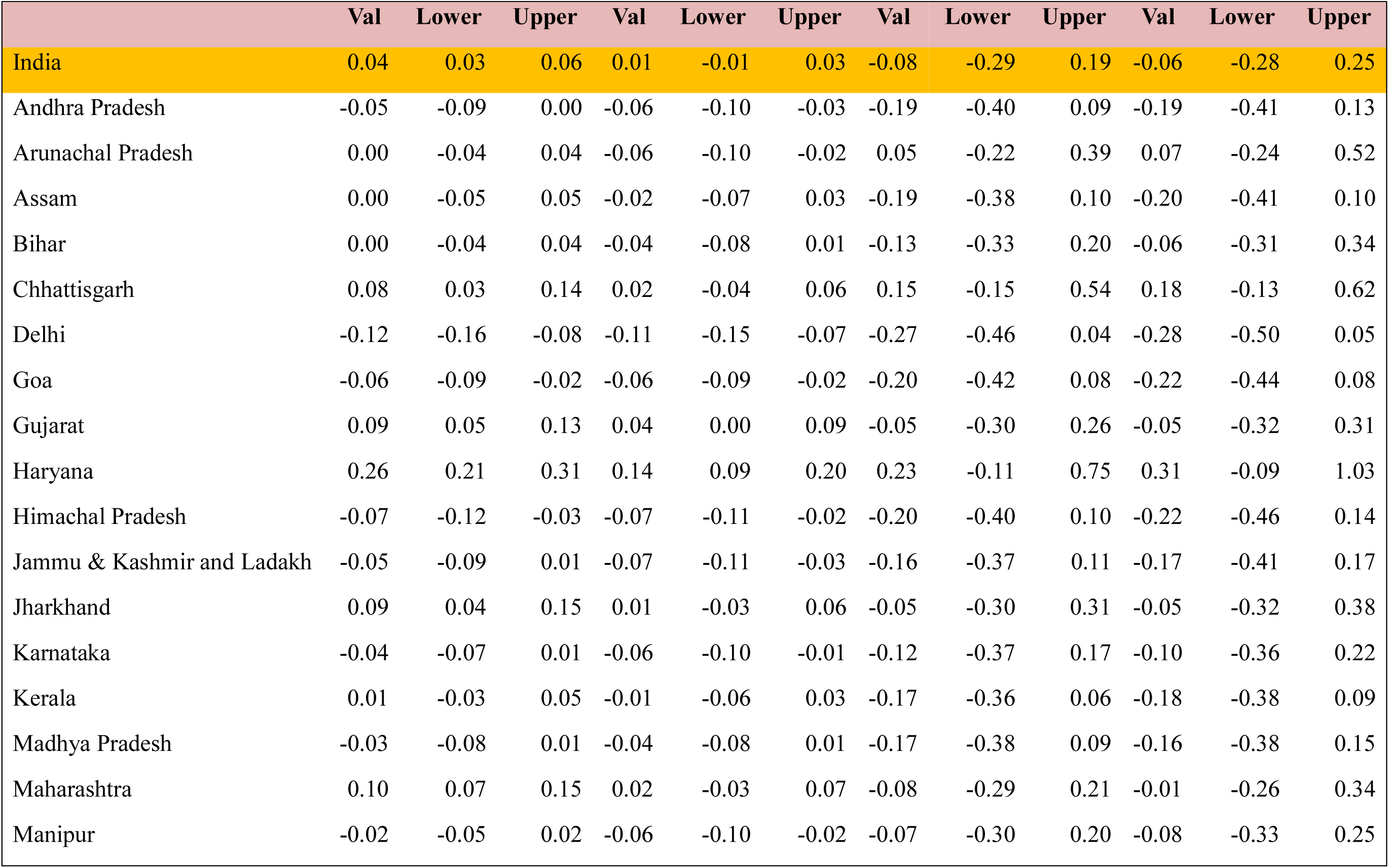
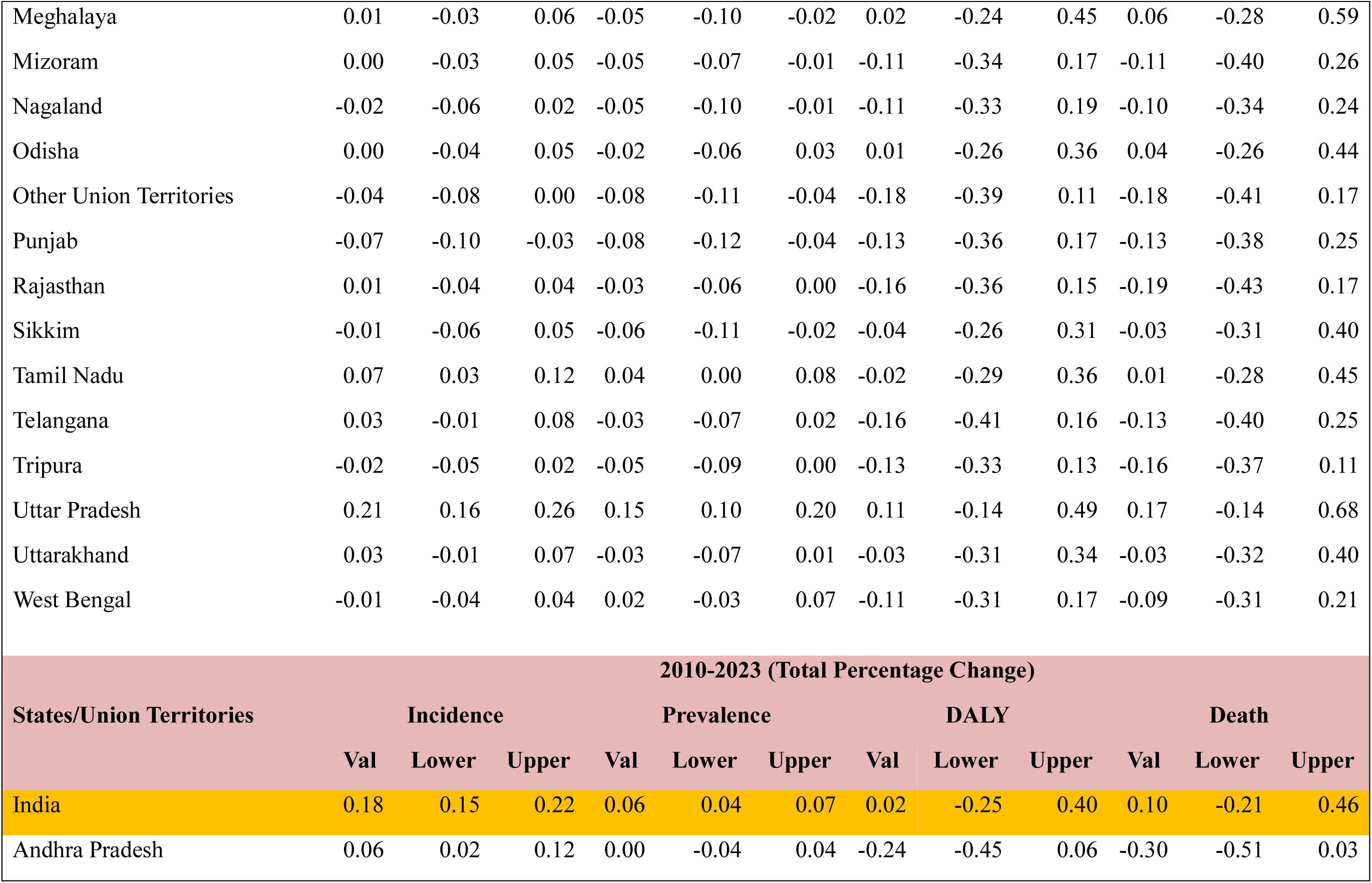
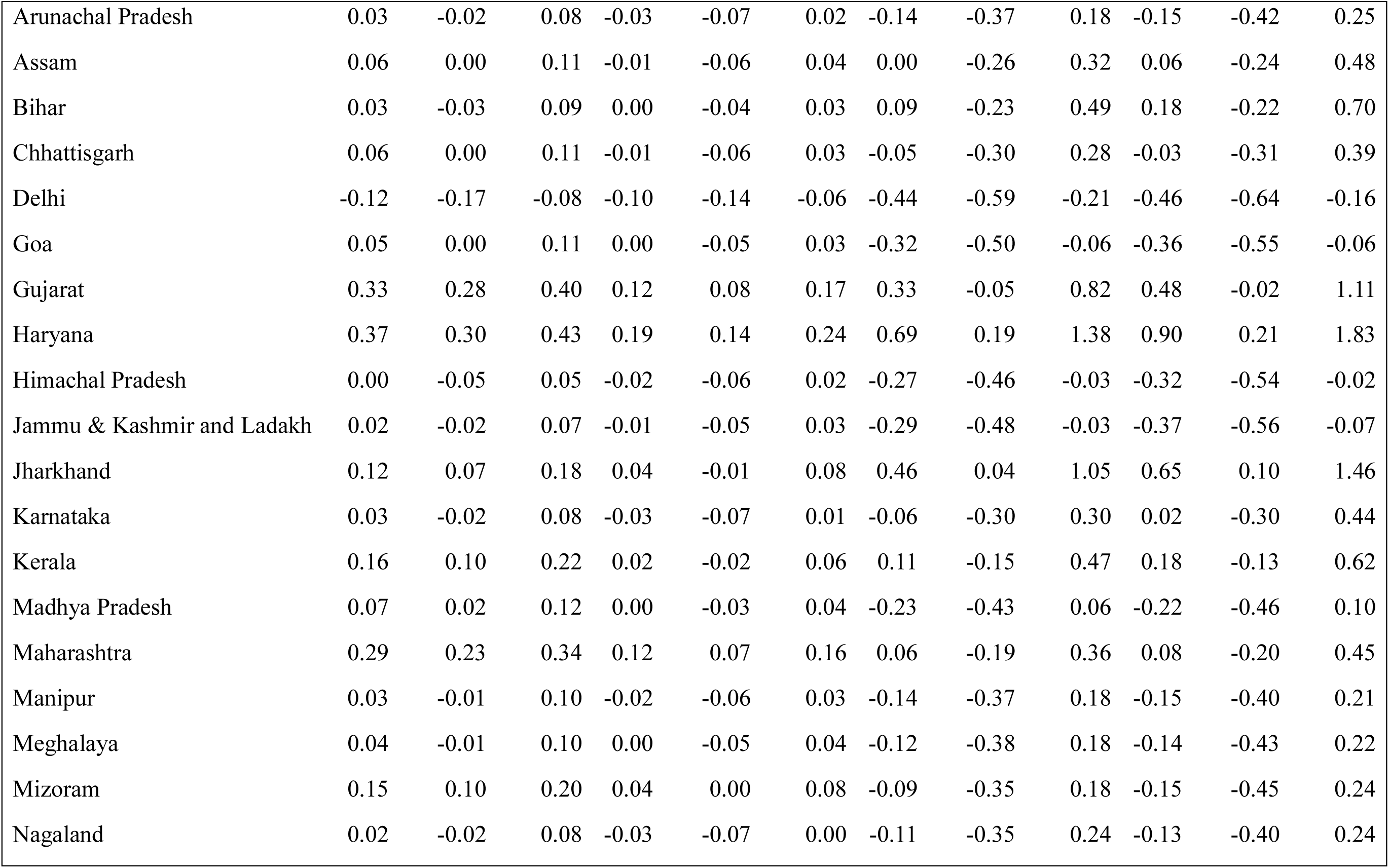
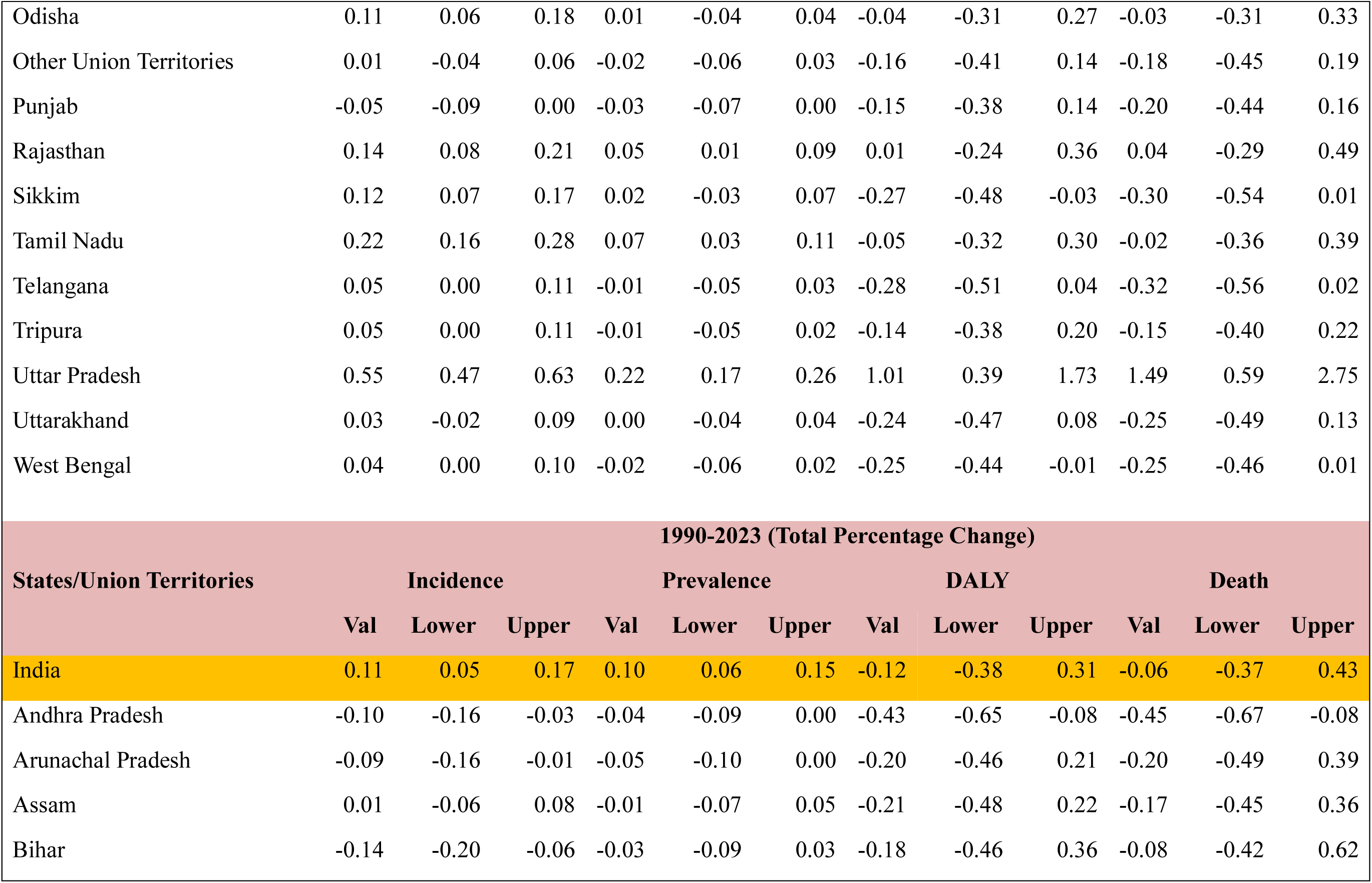
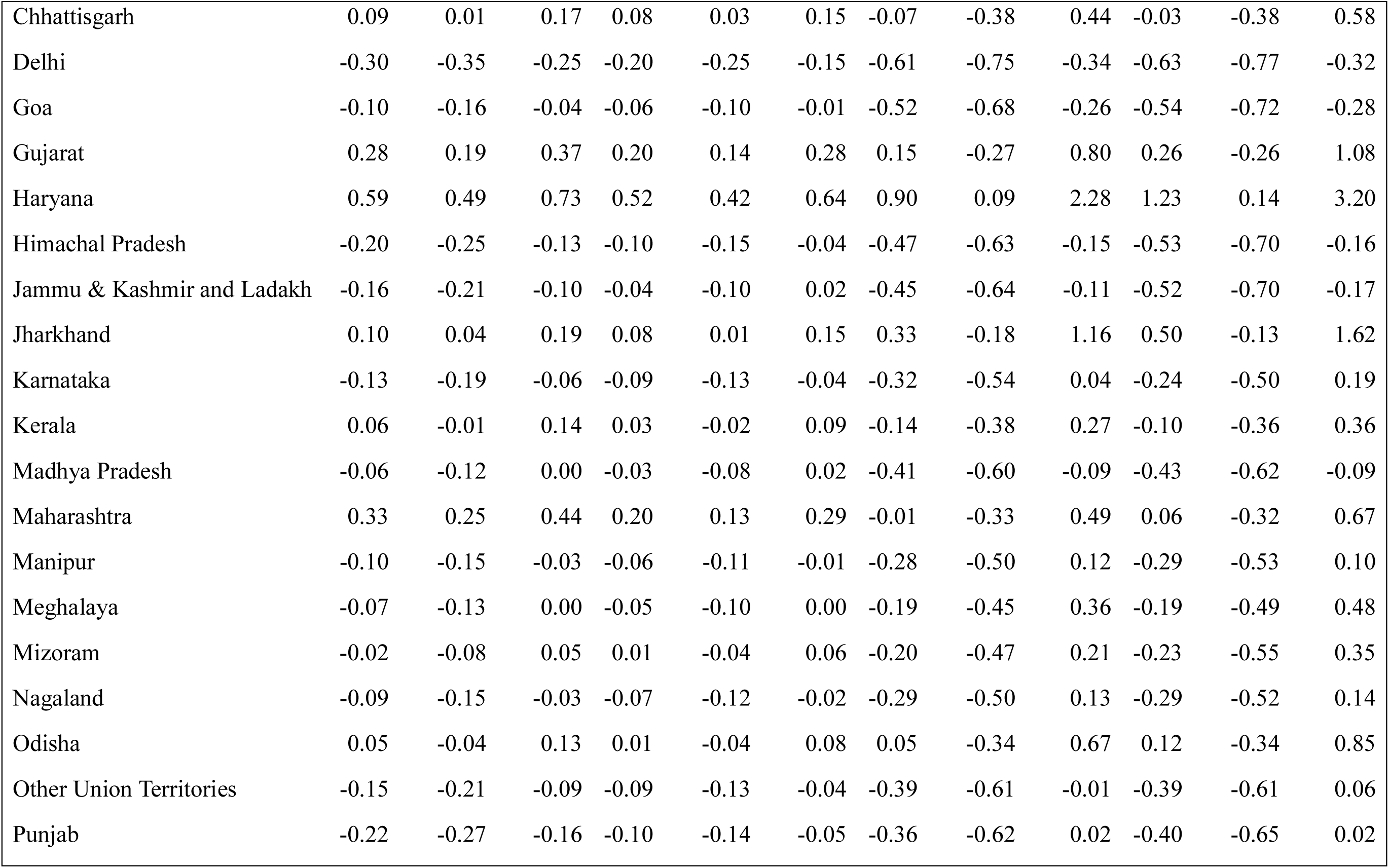
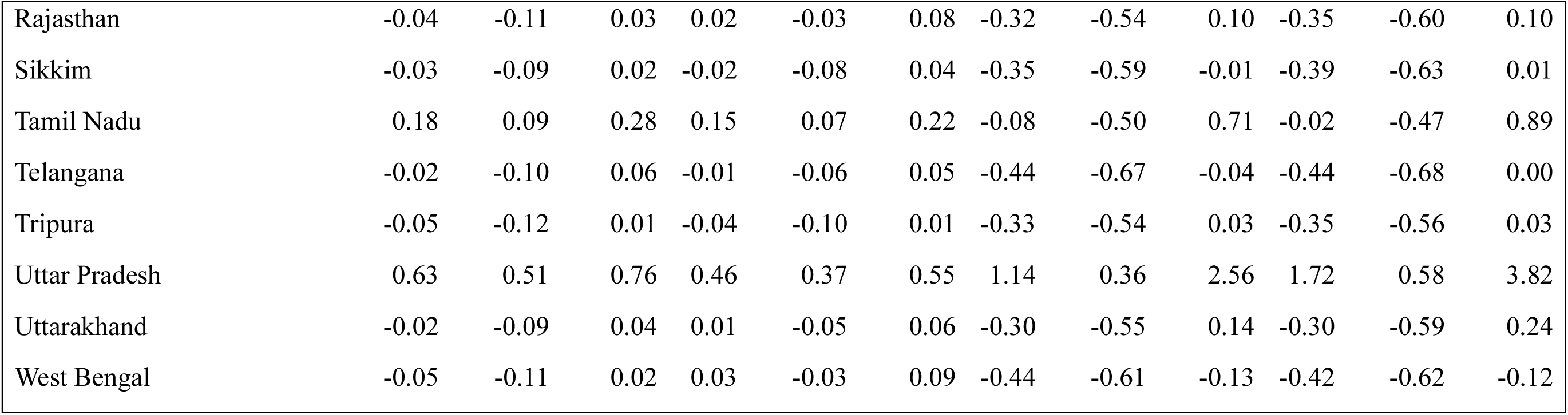
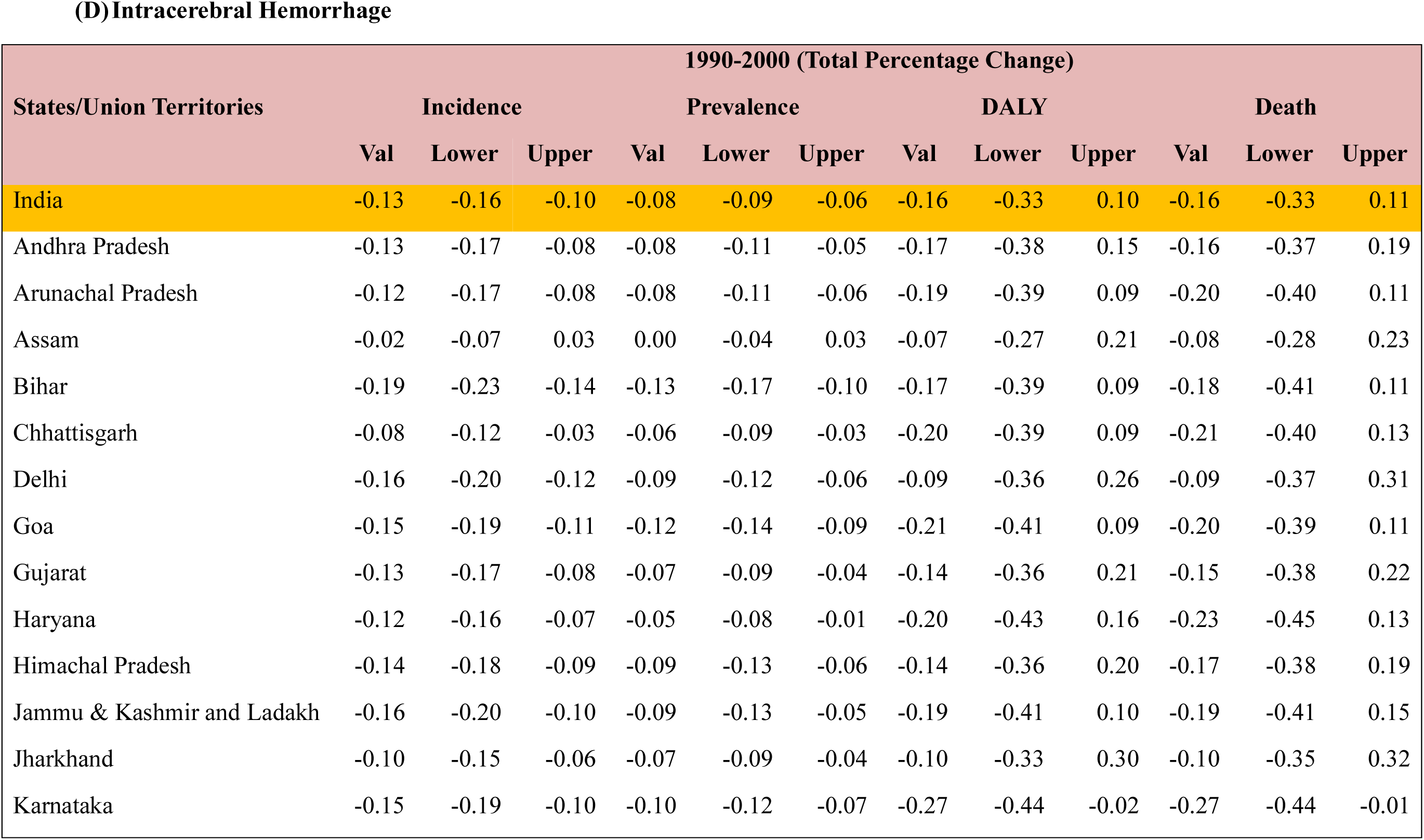
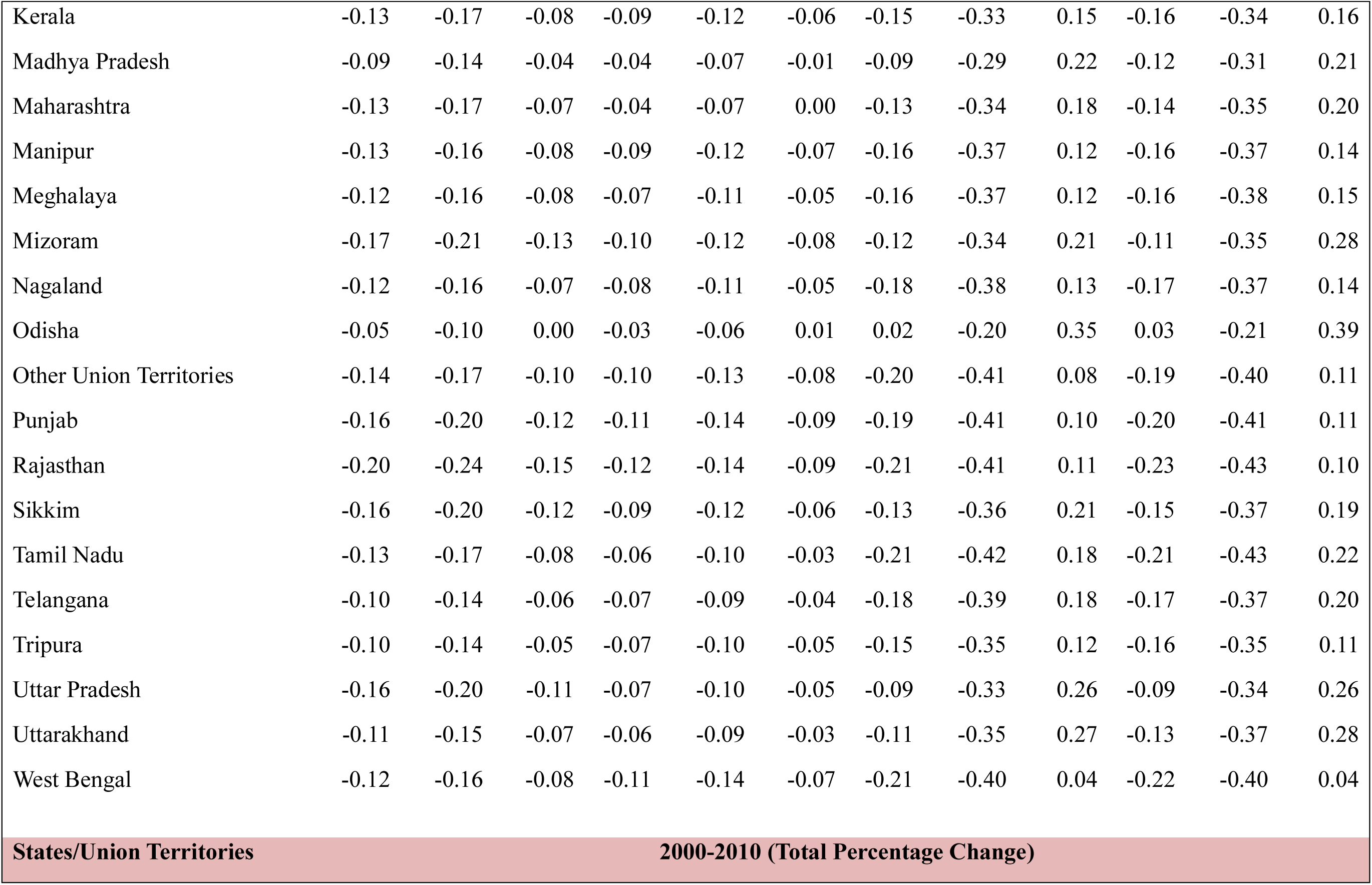
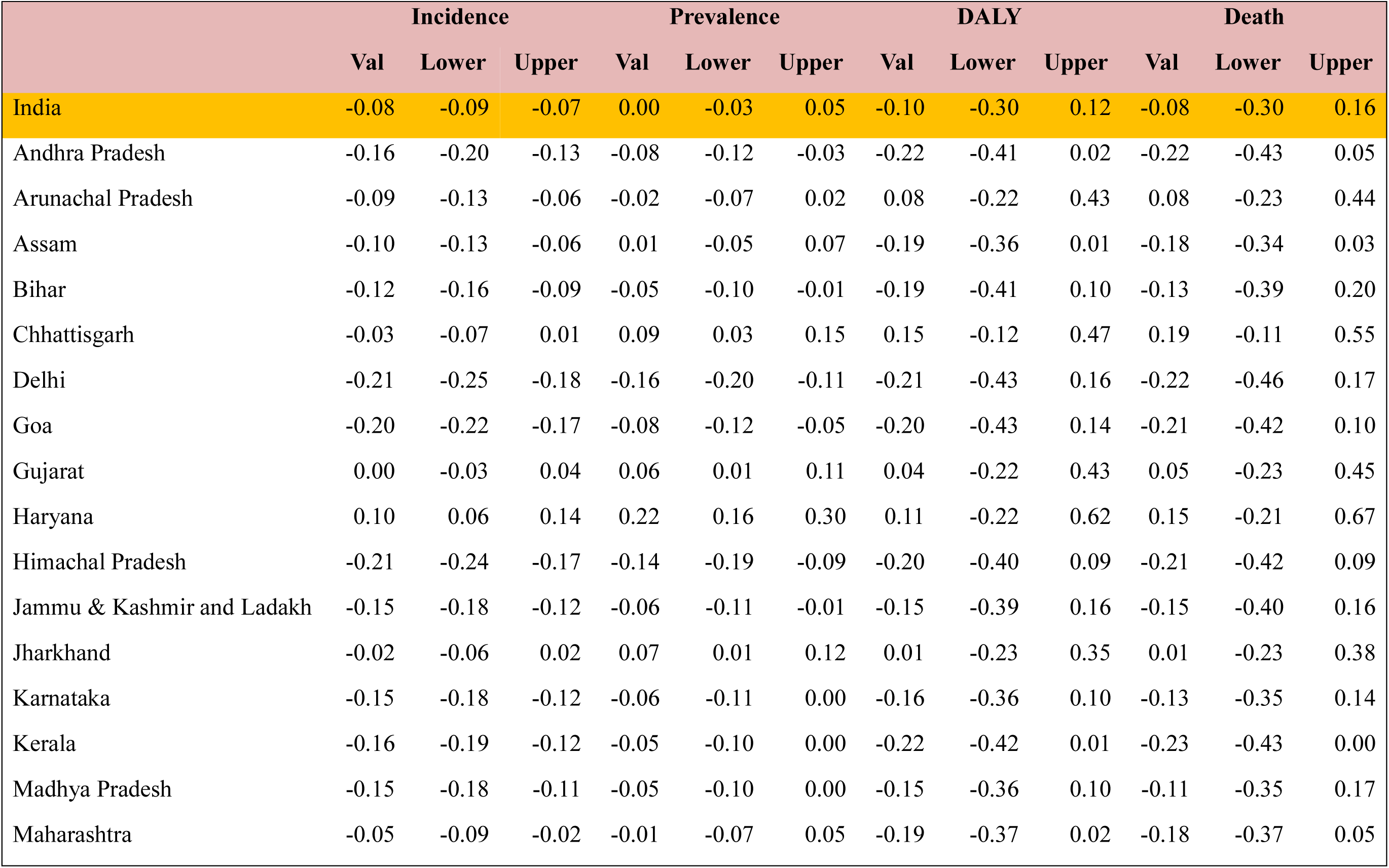
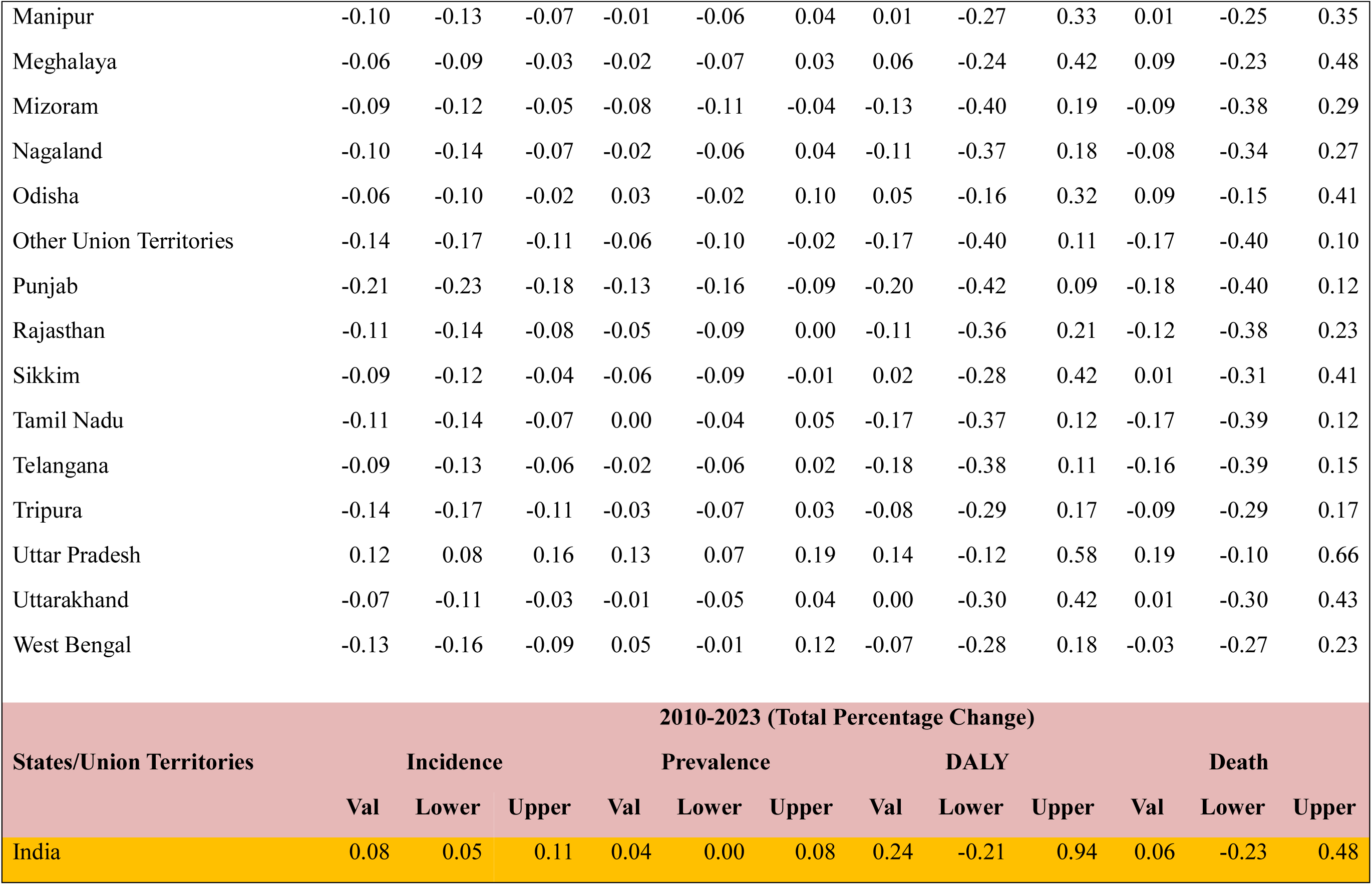
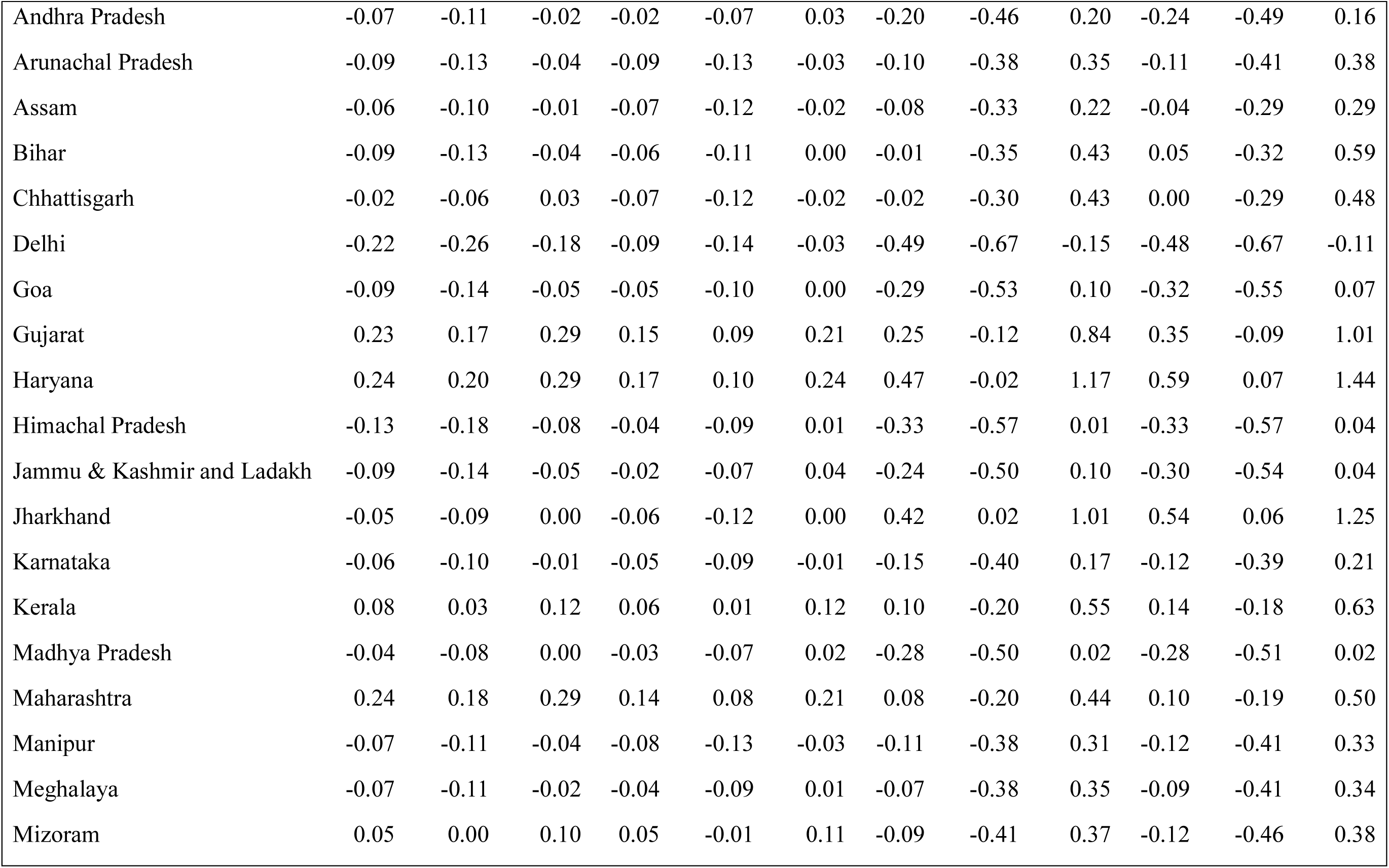
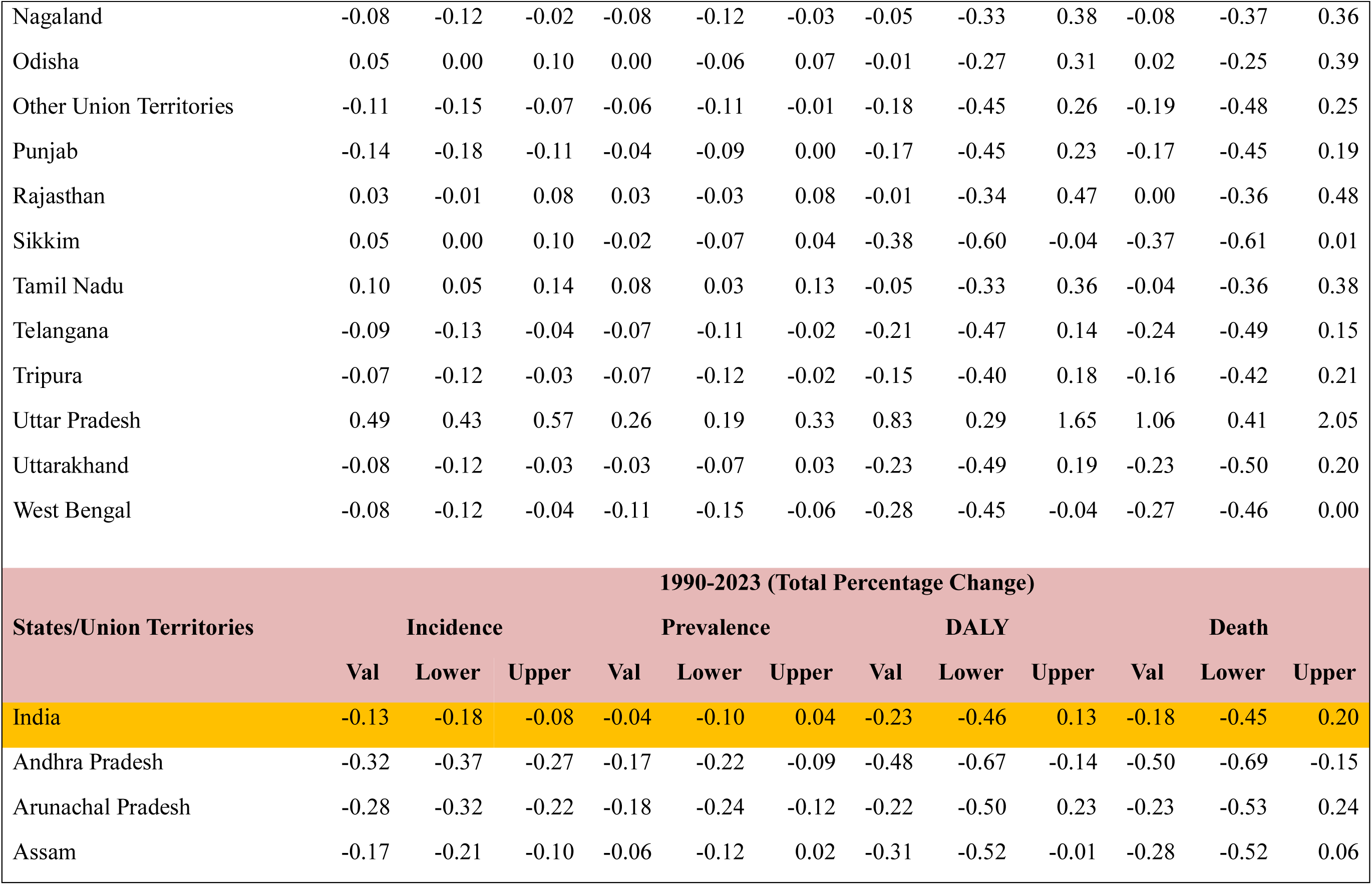
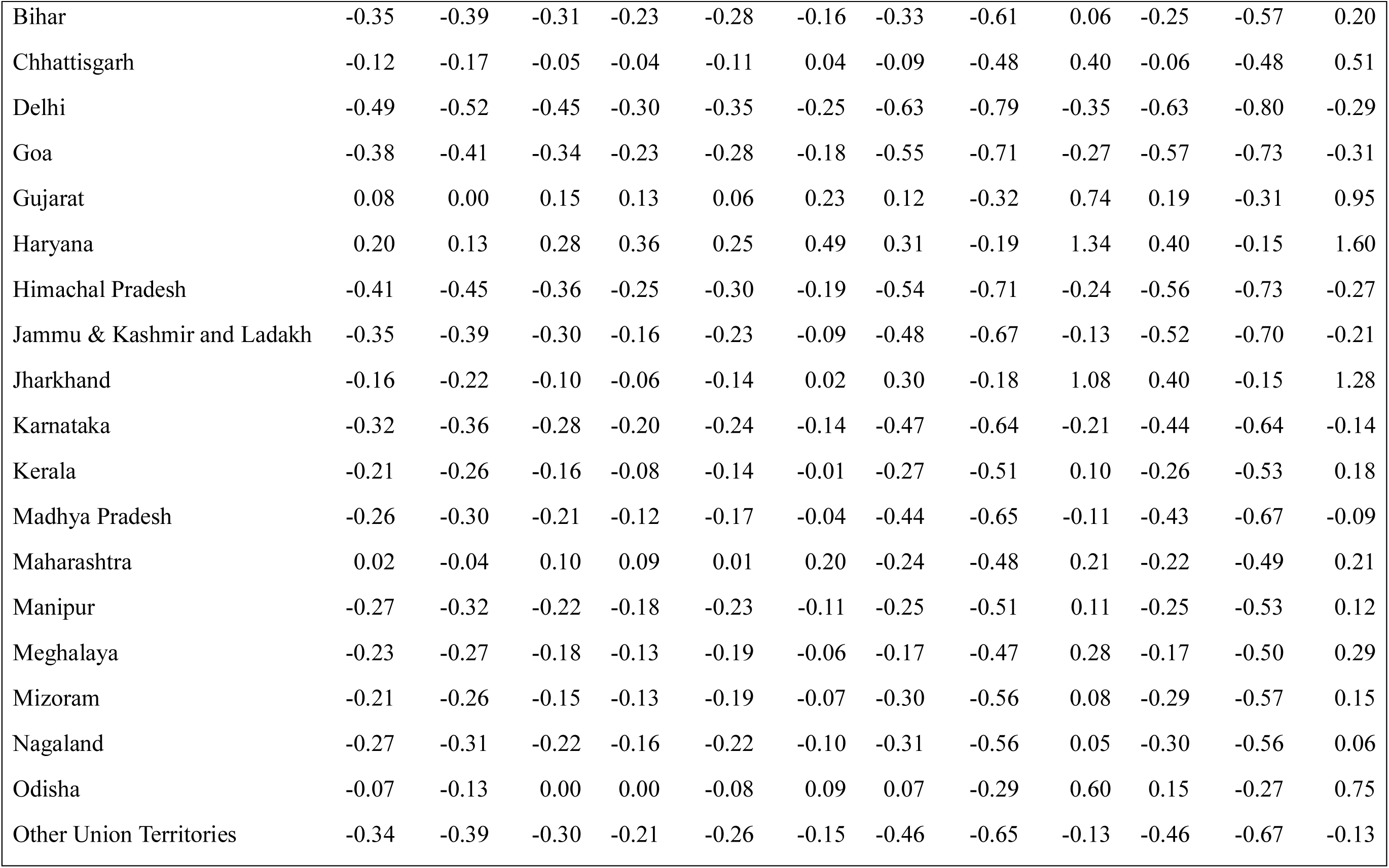
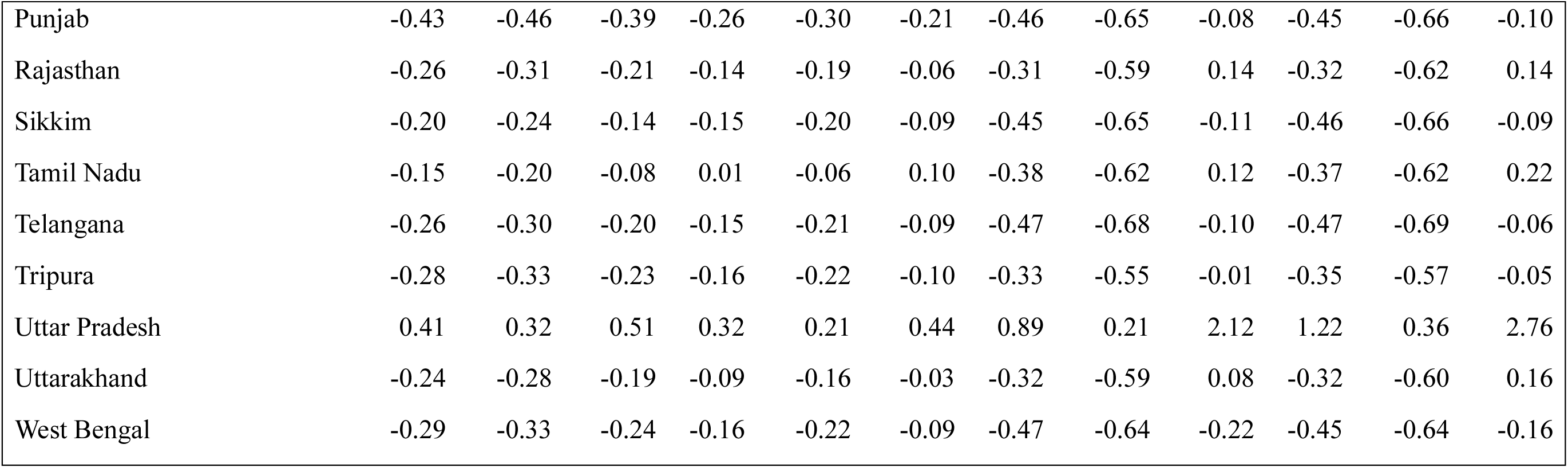
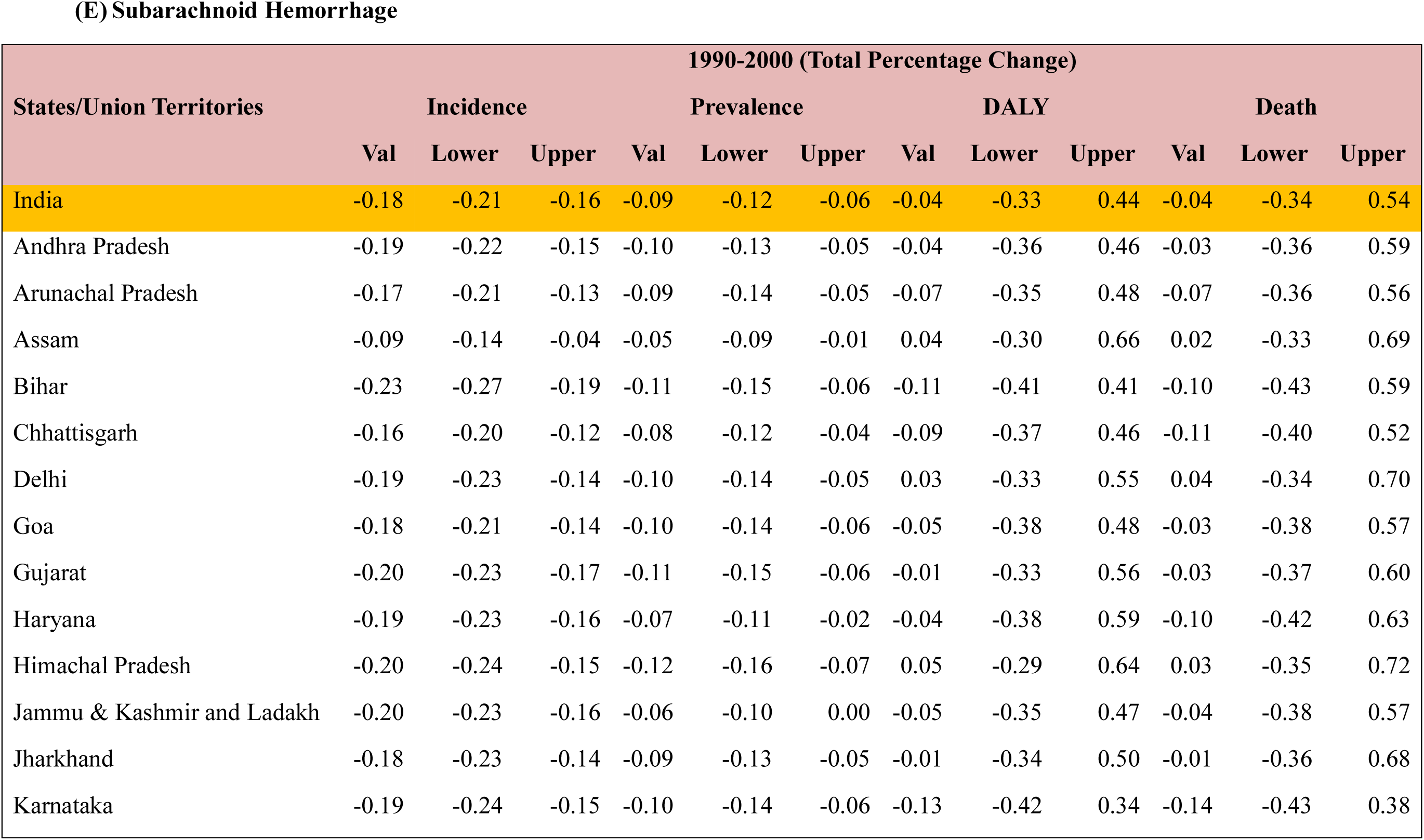
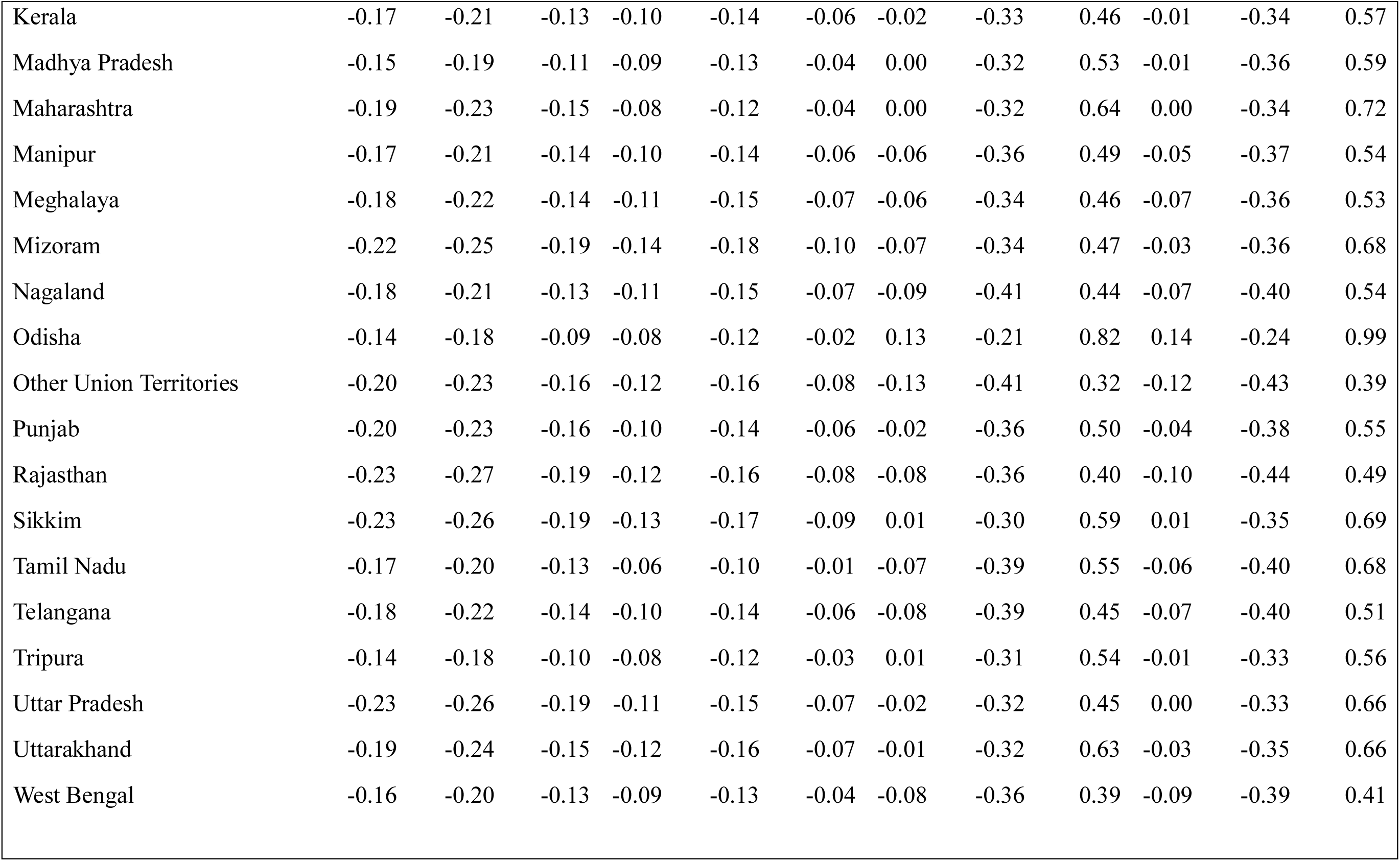
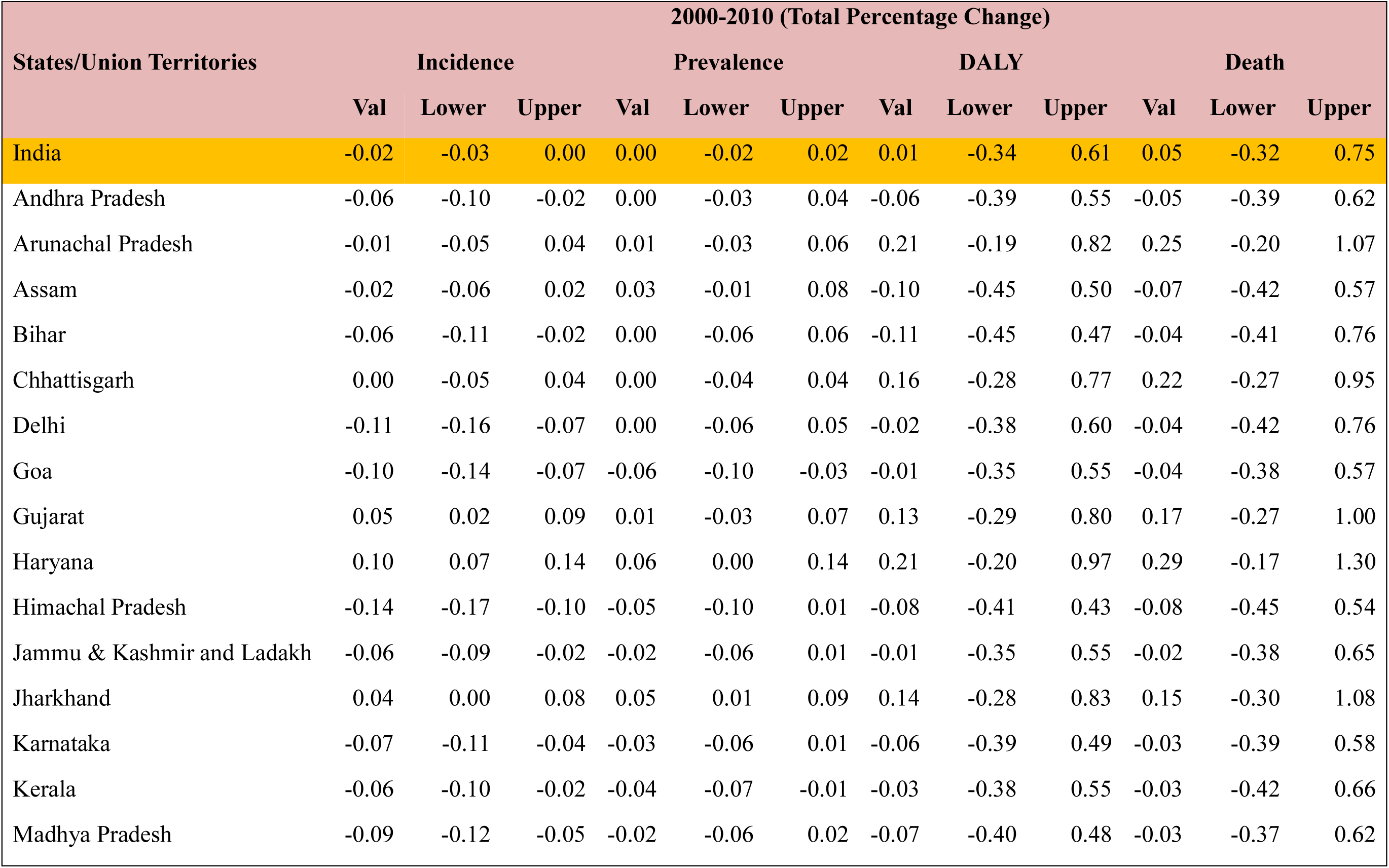
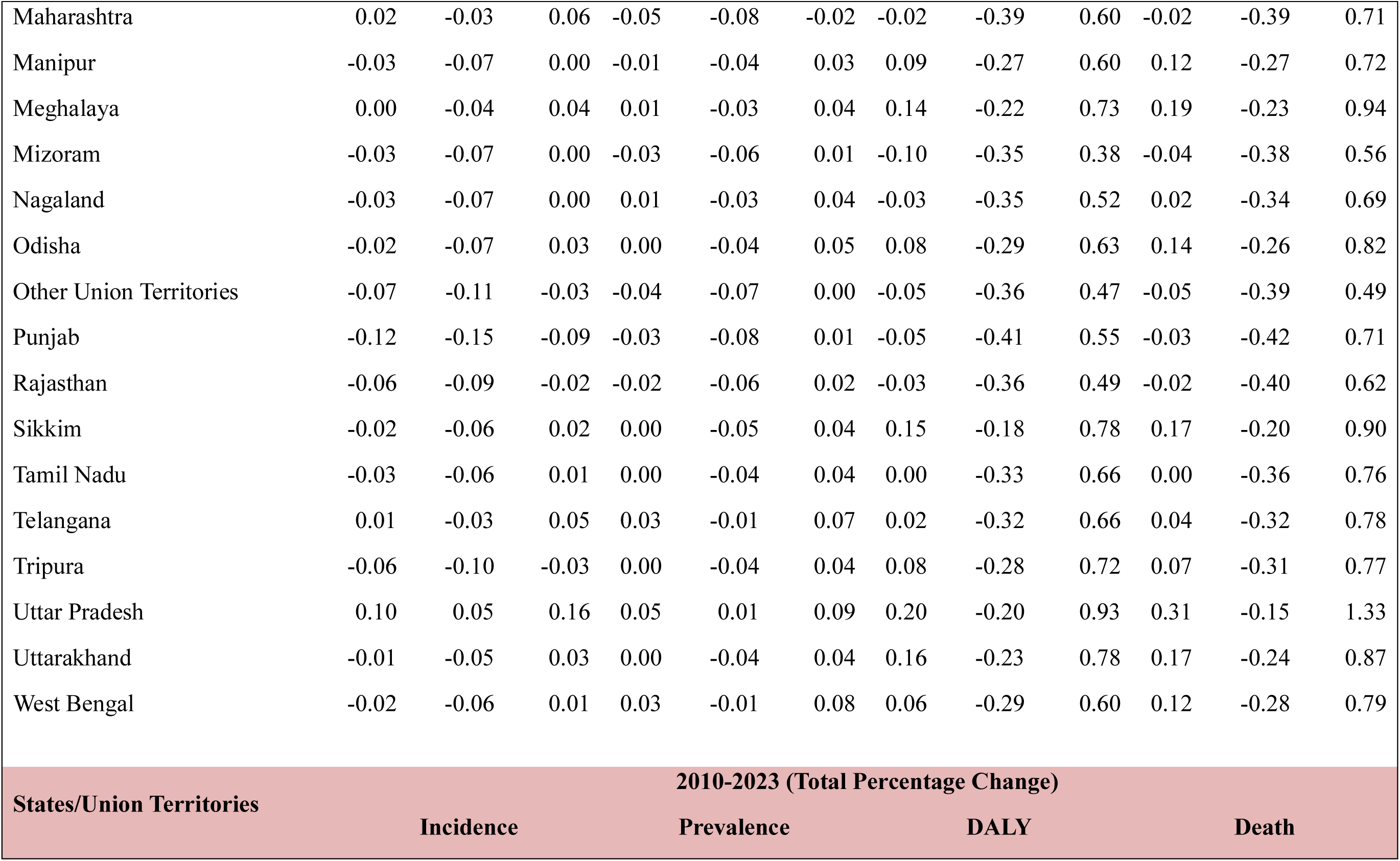
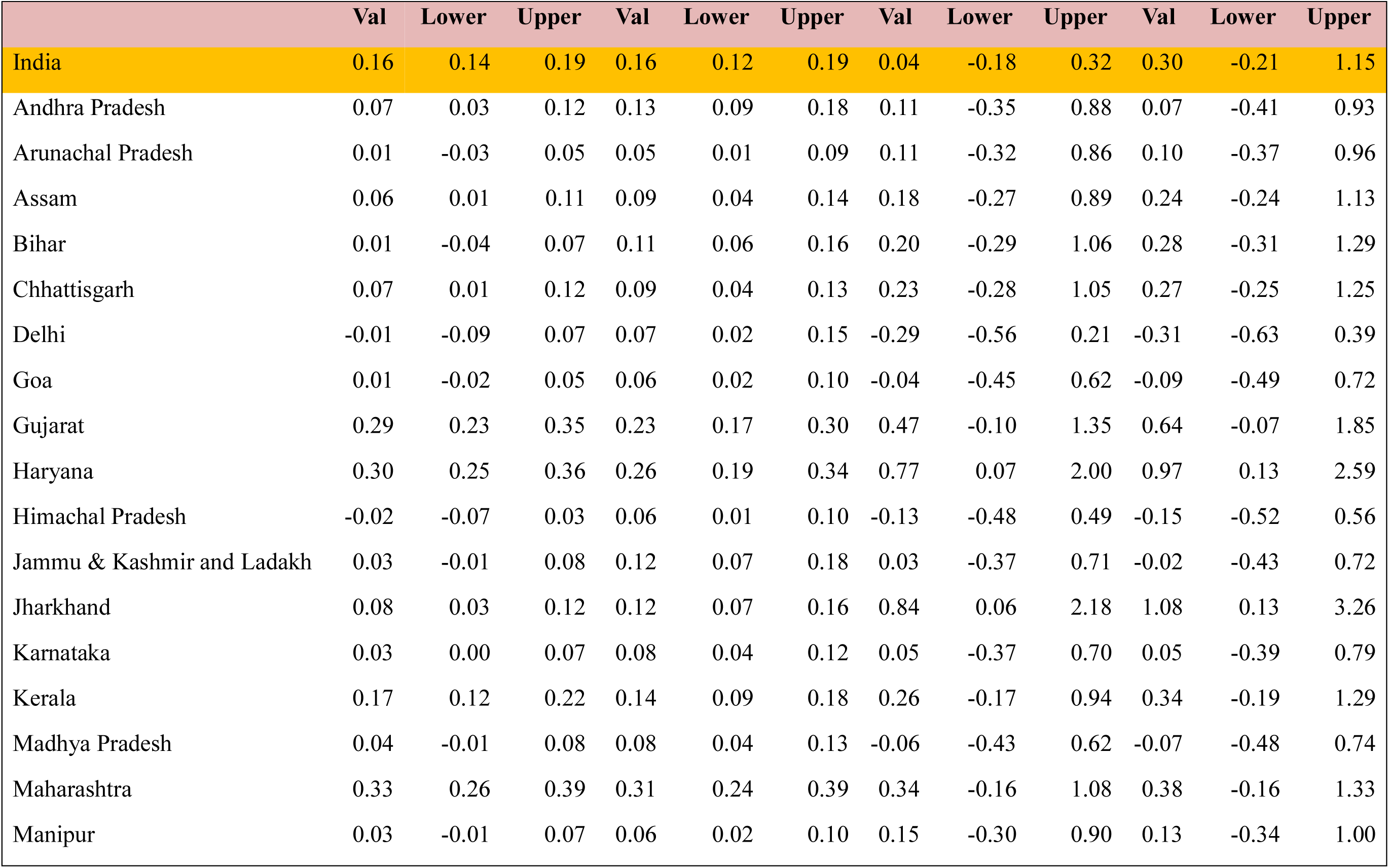
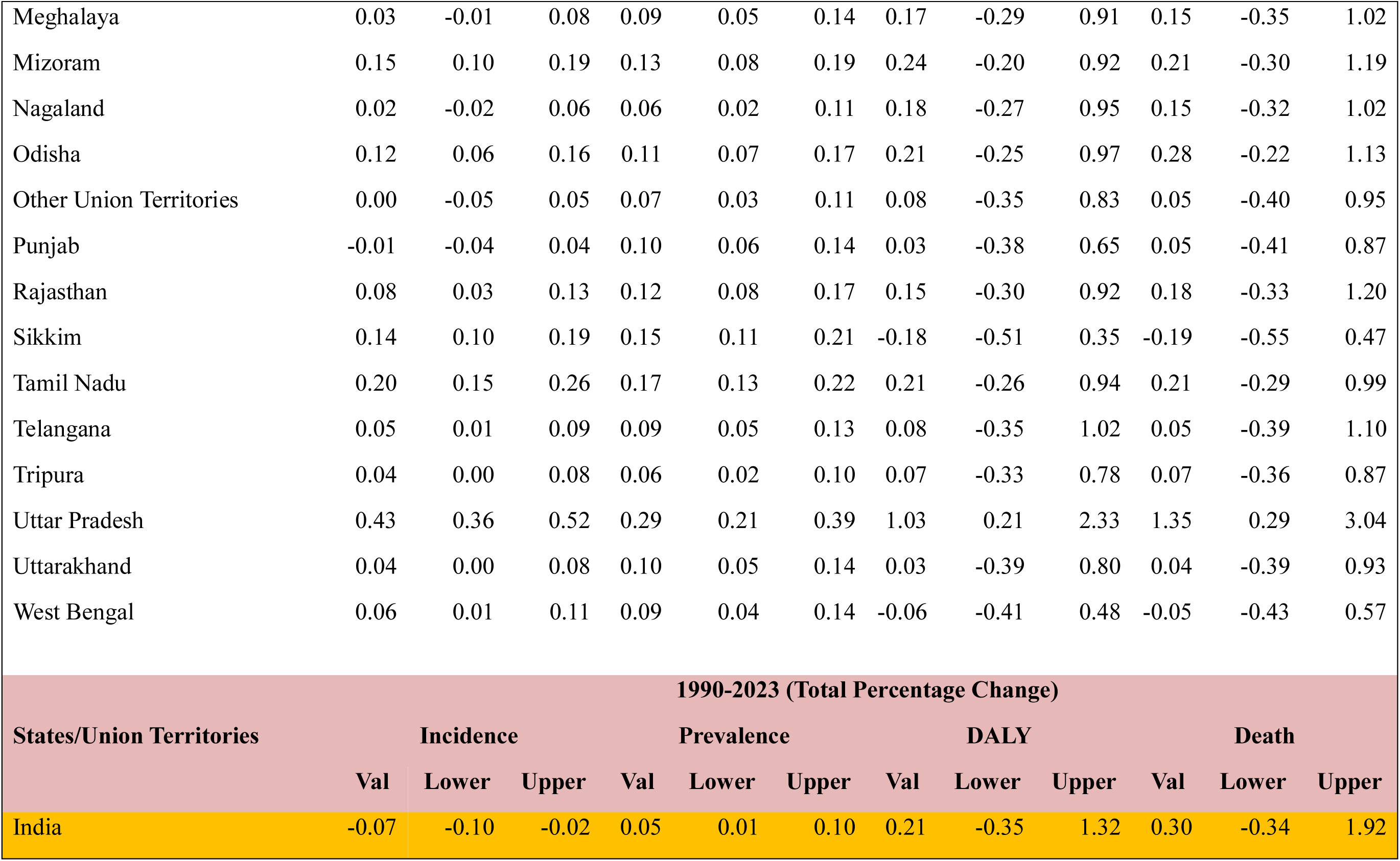
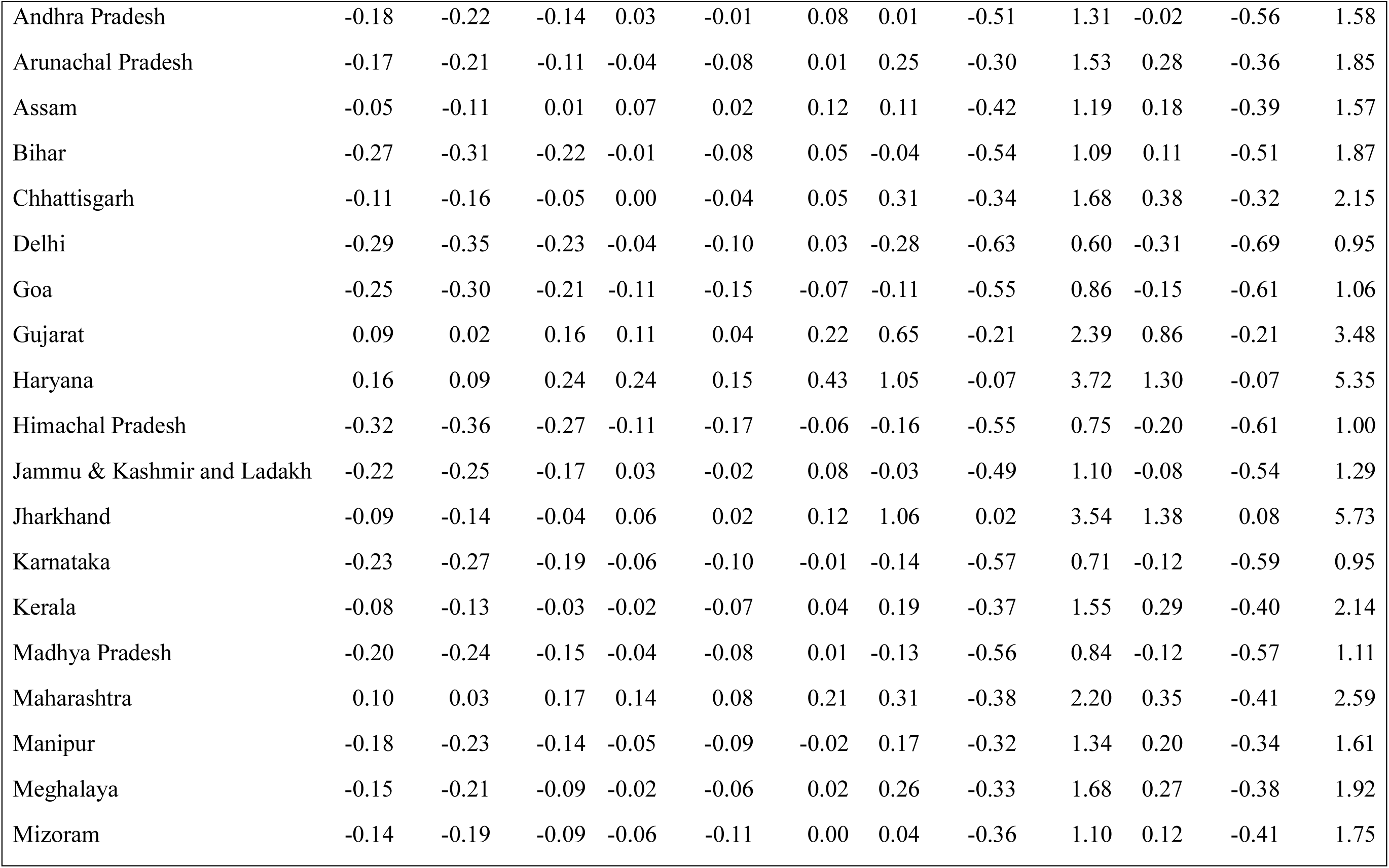
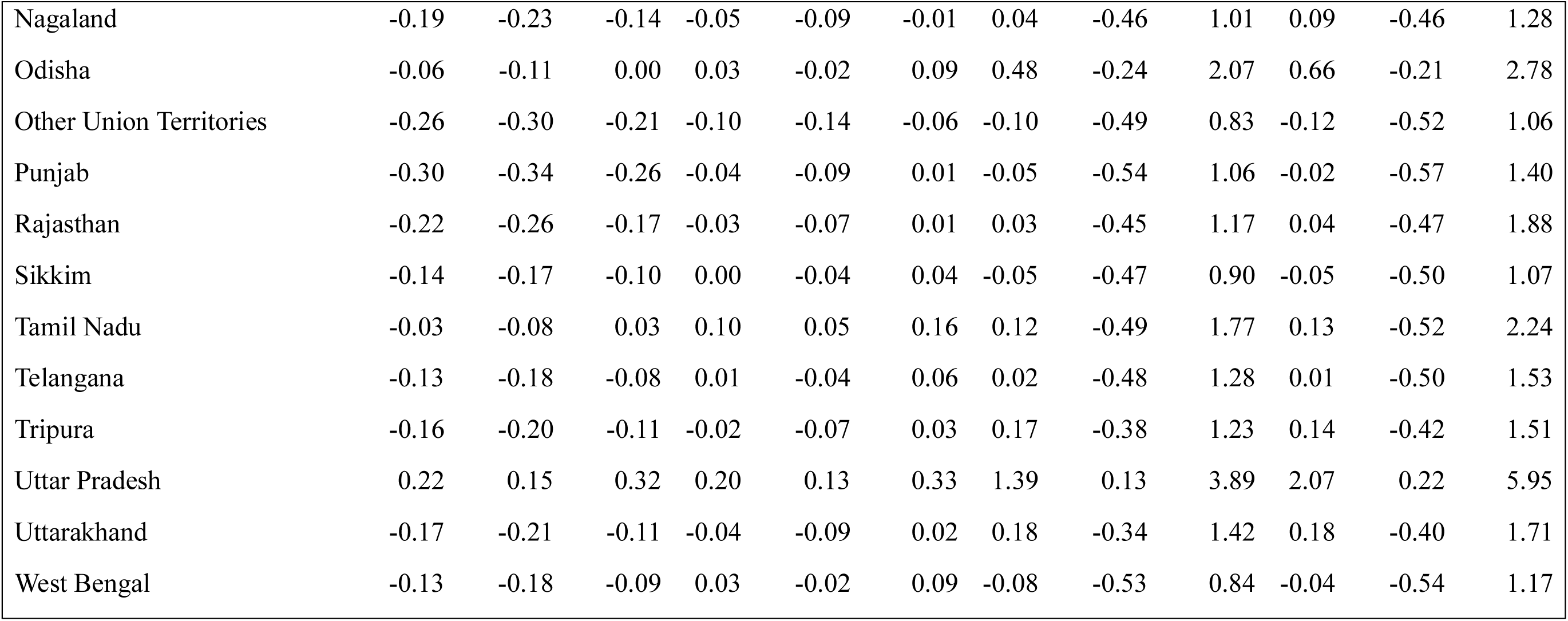
**(A-E):** (A) Total incident cases of stroke and its subtypes decade-wise from 1990 to 2023. Total Percentage Change of Incident Cases, Prevalent Cases, Disability-Adjusted Life Years (DALYs), and Deaths of (B) Overall Stroke, (C) Ischemic Stroke, (D) Intracerebral Hemorrhage, and (E) Subarachnoid Hemorrhage decade-wise from 1990-2000, 2000-2010, 2010-2023, and the overall 1990-2023 period

### Temporal Trends in Stroke Burden (1990–2023)

Between 1990 and 2023, the absolute number of stroke cases in India increased substantially, with incident cases rising from approximately 0.73 million (95% CI: 0.65–0.82 million) to 1.87 million (95% CI: 1.67–2.10 million). Despite this increase in absolute numbers, the age-standardized incidence rate remained largely unchanged over the study period, with a percentage change of 0.0% (95% CI: –0.04 to 0.05), indicating a stable underlying stroke risk after accounting for population structure.

Similarly, the number of stroke-related deaths increased in absolute terms from about 0.42 million (95% CI: 0.32–0.53 million) in 1990 to 1.03 million (95% CI: 0.83–1.26 million) in 2023. However, the age-standardized mortality rate declined slightly, with a percentage change of –0.11% (95% CI: –0.36 to 0.20), suggesting modest improvements in prevention, early detection, and clinical management.

In contrast, stroke prevalence showed a small increase, with age-standardized rates rising by 0.06% (95% CI: 0.03 to 0.10), reflecting improved survival and a growing population living with stroke. Meanwhile, age-standardized DALY rates declined modestly (–0.17% [95% CI: –0.38 to 0.12]), indicating some progress in reducing overall health loss, possibly due to better acute care and rehabilitation services.

Overall, while age-standardized rates have remained stable or slightly improved, the absolute burden of stroke has increased considerably, largely driven by population growth and aging (**Table 1 and Figure 1**).

**Figure 1:**
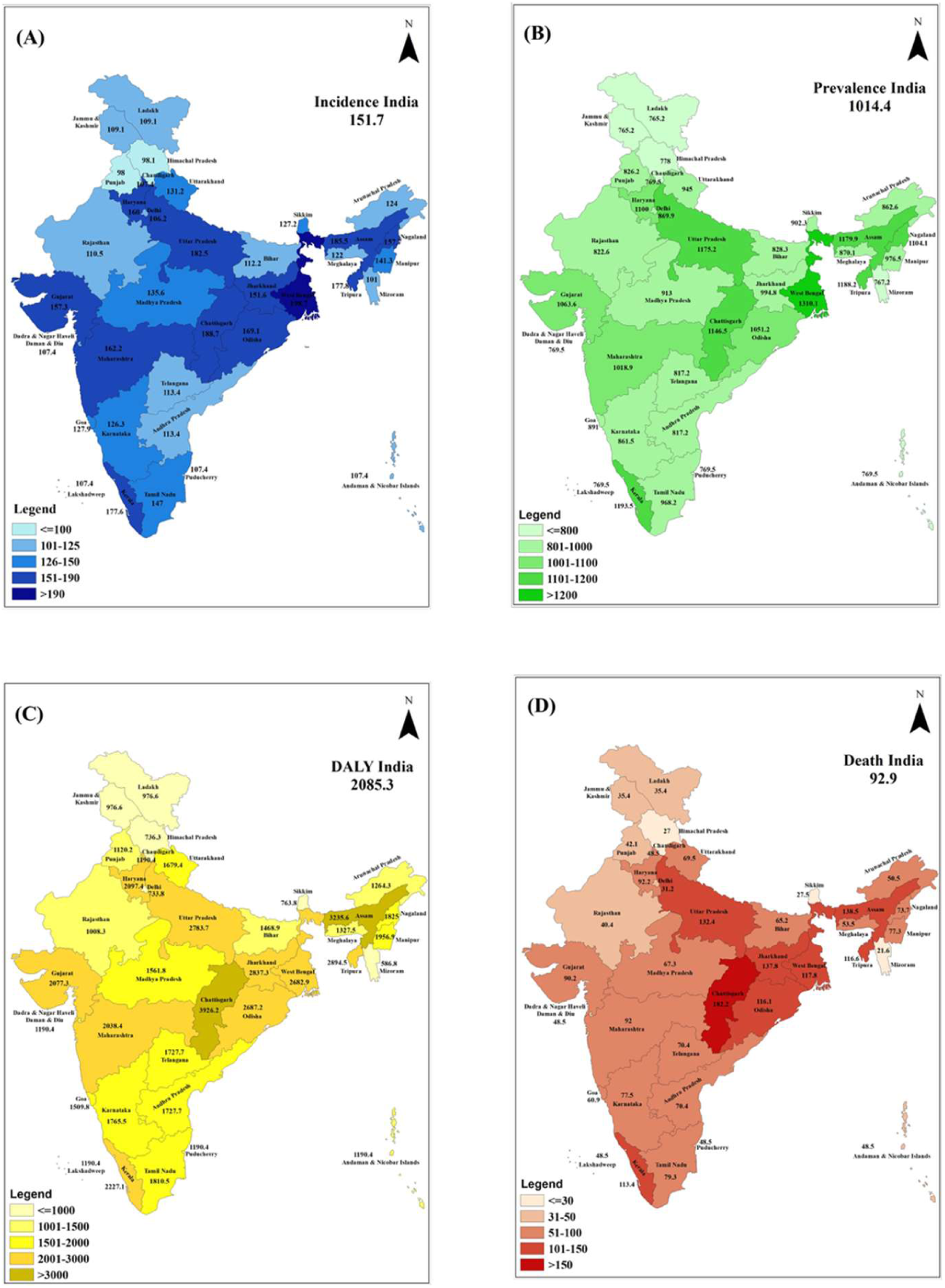
Percentage Change of Stroke Burden Across Indian States from 1990 to 2023; DALY, disability adjusted life-year. (A) Percent Change of Incidence Rate (B) Percent Change of Prevalence Rate (C) Percent Change of DALYs Rate (D) Percent Change of Death Rate

### Decade-wise Temporal Changes in Stroke Burden (1990–2023)

Over the period from 1990 to 2023, temporal patterns of stroke burden varied across subtypes in India. The incidence of ischaemic stroke increased markedly in absolute terms, rising from approximately 382,691 cases (95% CI: 315,908–449,948) in 1990 to 1,092,415 cases (95% CI: 940,855–1,278,944) in 2023. However, the age-standardized percentage change suggests only a modest overall increase. In contrast, although the number of intracerebral hemorrhage cases increased from about 290,024 (95% CI: 238,849–341,500) to 649,269 (95% CI: 544,109–753,887) over the same period, the age-standardized incidence declined, indicating a reduction in the underlying risk after accounting for population structure. A similar pattern was observed for subarachnoid hemorrhage, where incident cases increased from 59,427 (95% CI: 49,892–70,181) in 1990 to 126,209 (95% CI: 108,345–144,835) in 2023, but with a slight decrease in age-standardized incidence rates. Overall, these findings suggest that while absolute numbers have increased across all stroke subtypes, the underlying risk has remained stable or declined modestly (Table 1 and Fig. 2).

**Figure 2:**
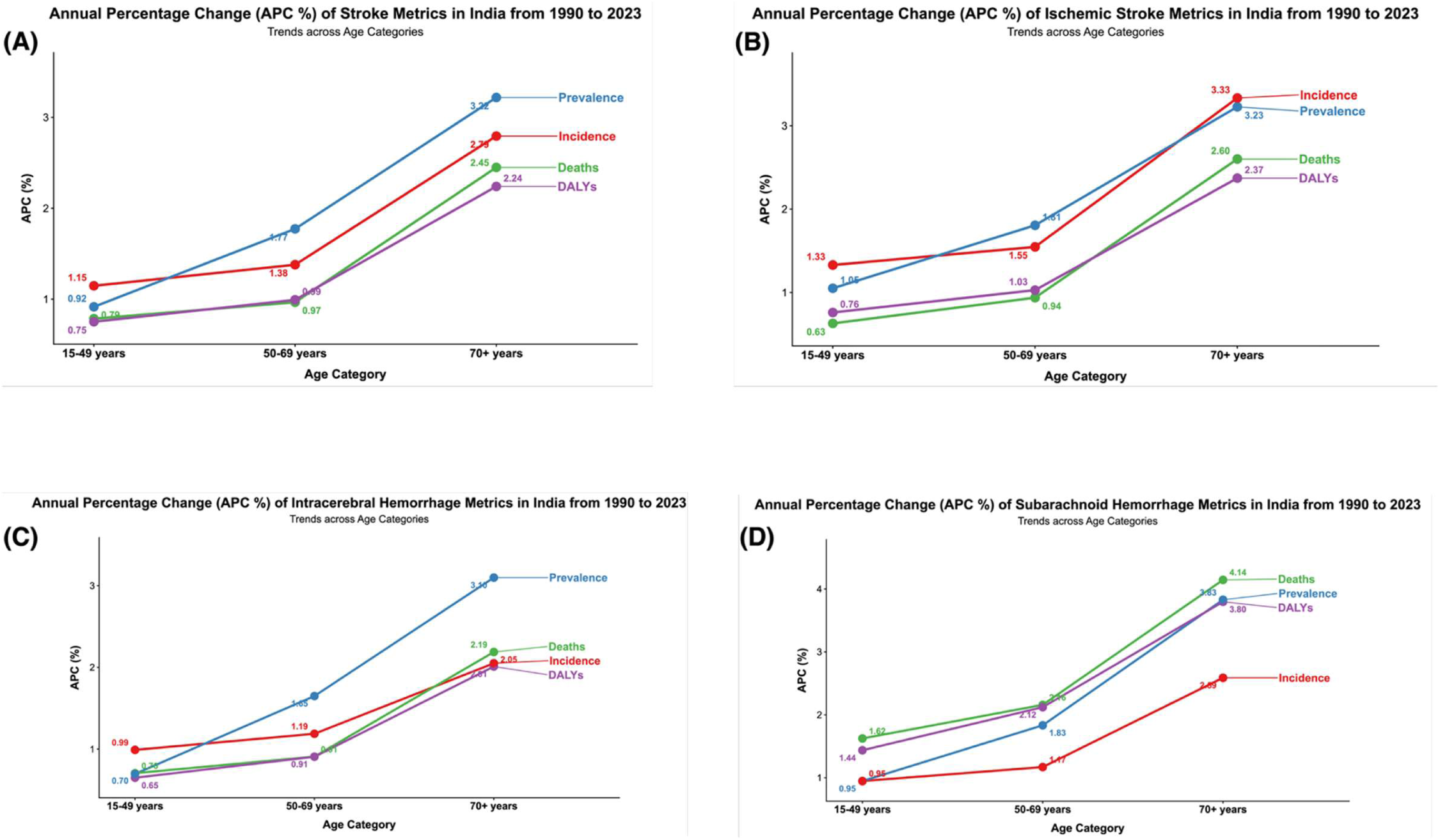
Trends of stroke onset across age categories from 1990 to 2023; Annual Percentage change (APC%) of Stroke (B), Annual Percentage change (APC%) of Ischemic Stroke (C), Annual Percentage change (APC%) of Intracerebral Hemorrhage (D), Annual Percentage change (APC%) of Subarachnoid Hemorrhage

### State-Level Variations in Stroke Burden

There was considerable geographic variation in stroke burden across Indian states. In 2023, the highest age-standardized stroke mortality rates were observed in Chhattisgarh (182.16 per 100,000; 95% CI: 133.87–230.89), Assam (138.45 per 100,000; 95% CI: 107.78–174.48), and Jharkhand (137.84 per 100,000; 95% CI: 105.26–173.27). In contrast, the lowest mortality rates were reported in Mizoram (21.62 per 100,000; 95% CI: 14.56–31.49), Himachal Pradesh (27.02 per 100,000; 95% CI: 18.69–38.03), and Sikkim (27.49 per 100,000; 95% CI: 19.12–37.87).

A similar pattern was observed for age-standardized DALY rates, which ranged widely from 3,926.20 per 100,000 (95% CI: 2,897.81–4,924.59) in Chhattisgarh to 586.83 per 100,000 (95% CI: 425.48–809.90) in Mizoram. These findings highlight substantial regional disparities in stroke burden across India, reflecting differences in risk factor distribution, healthcare access, and socioeconomic conditions (**Fig. 3**).

**Figure 3:**
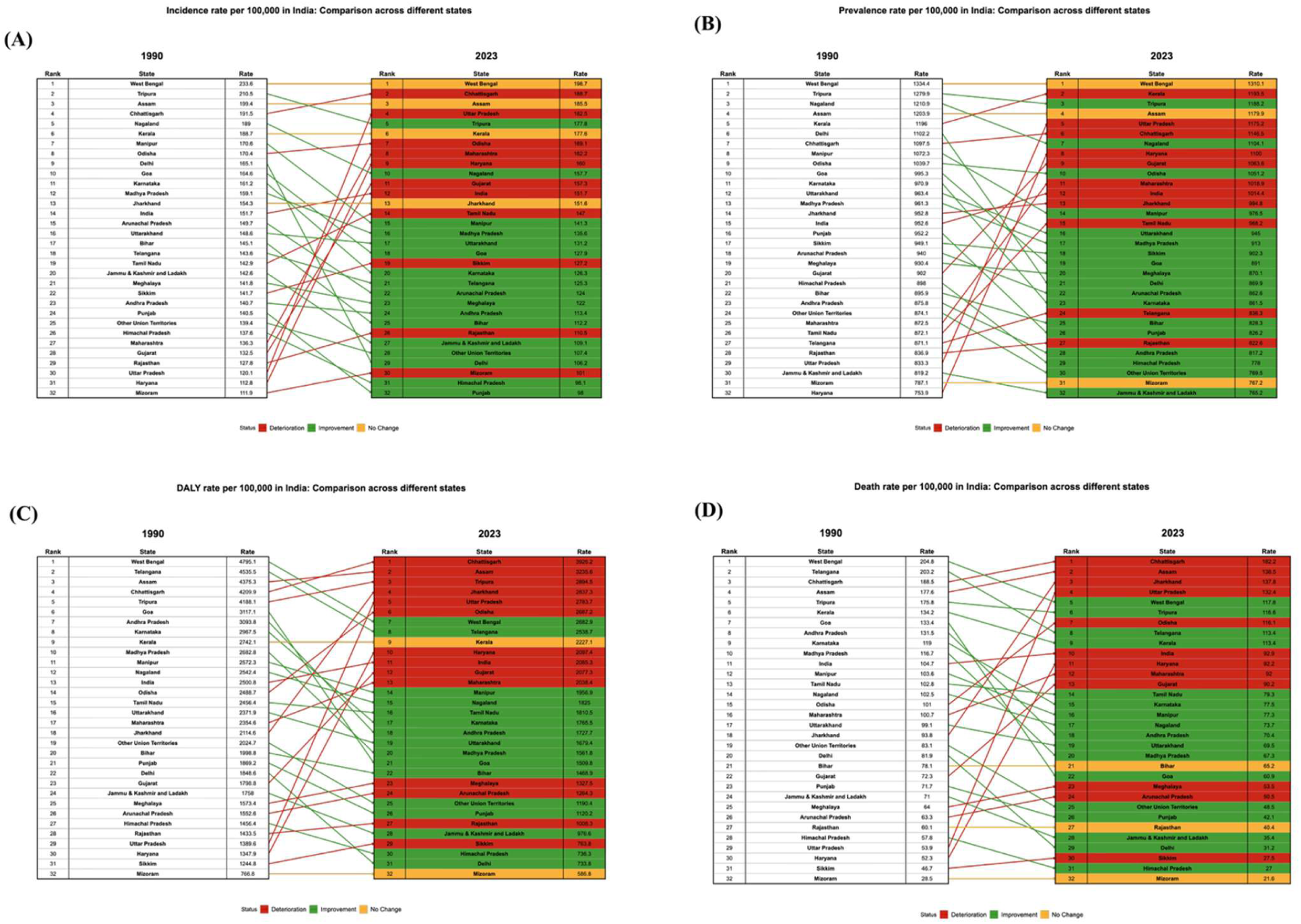
Change in the rank of stroke in terms of age-standardized rate of (A) Incidence, (B) Prevalence, (C) DALYs, (D) Death in India, 1990-2023; DALYs=disability-adjusted life-years

### Risk Factor Attribution

A significant proportion of stroke burden in India was attributable to modifiable risk factors. In 2023, high systolic blood pressure remained the leading contributor, accounting for 0.546% (95% CI: 0.377–0.686) of stroke deaths and 0.534% (95% CI: 0.368–0.679) of stroke DALYs. Other important contributors included dietary risks (0.196%; 95% CI: 0.049–0.330), ambient particulate matter pollution (0.182%; 95% CI: 0.116–0.253), tobacco use (0.089%; 95% CI: 0.046–0.145), high fasting plasma glucose (0.083%; 95% CI: 0.059–0.123), and high body-mass index (0.015%; 95% CI: 0.0001–0.036). These findings underscore the dominant role of modifiable behavioral, metabolic, and environmental factors in shaping stroke burden (**Fig. 4-6**).

**Figure 4:**
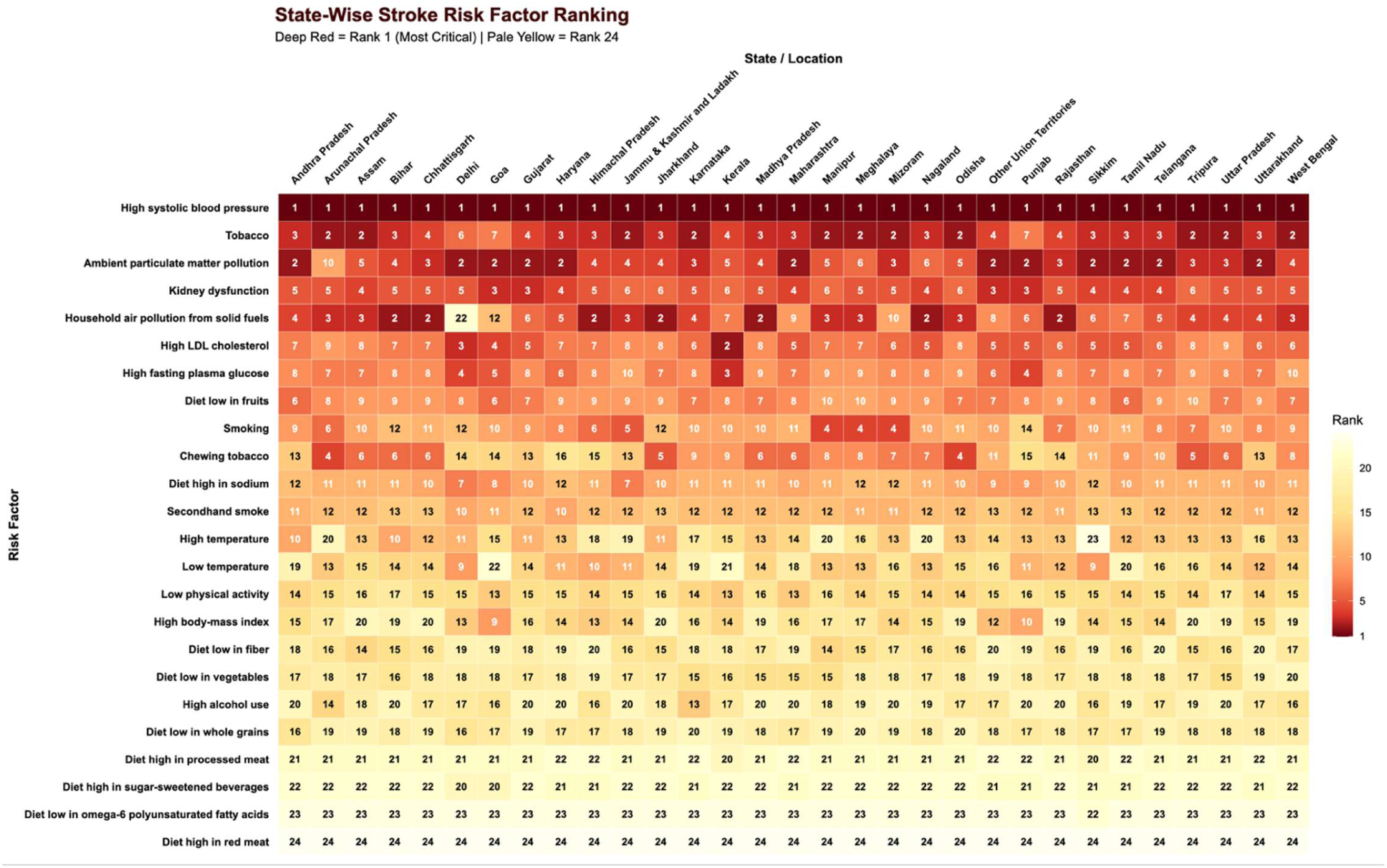
Ranking of age-standardized stroke death attributable to risk factors for both sexes, state-wise distribution for the year 2023

**Figure 5:**
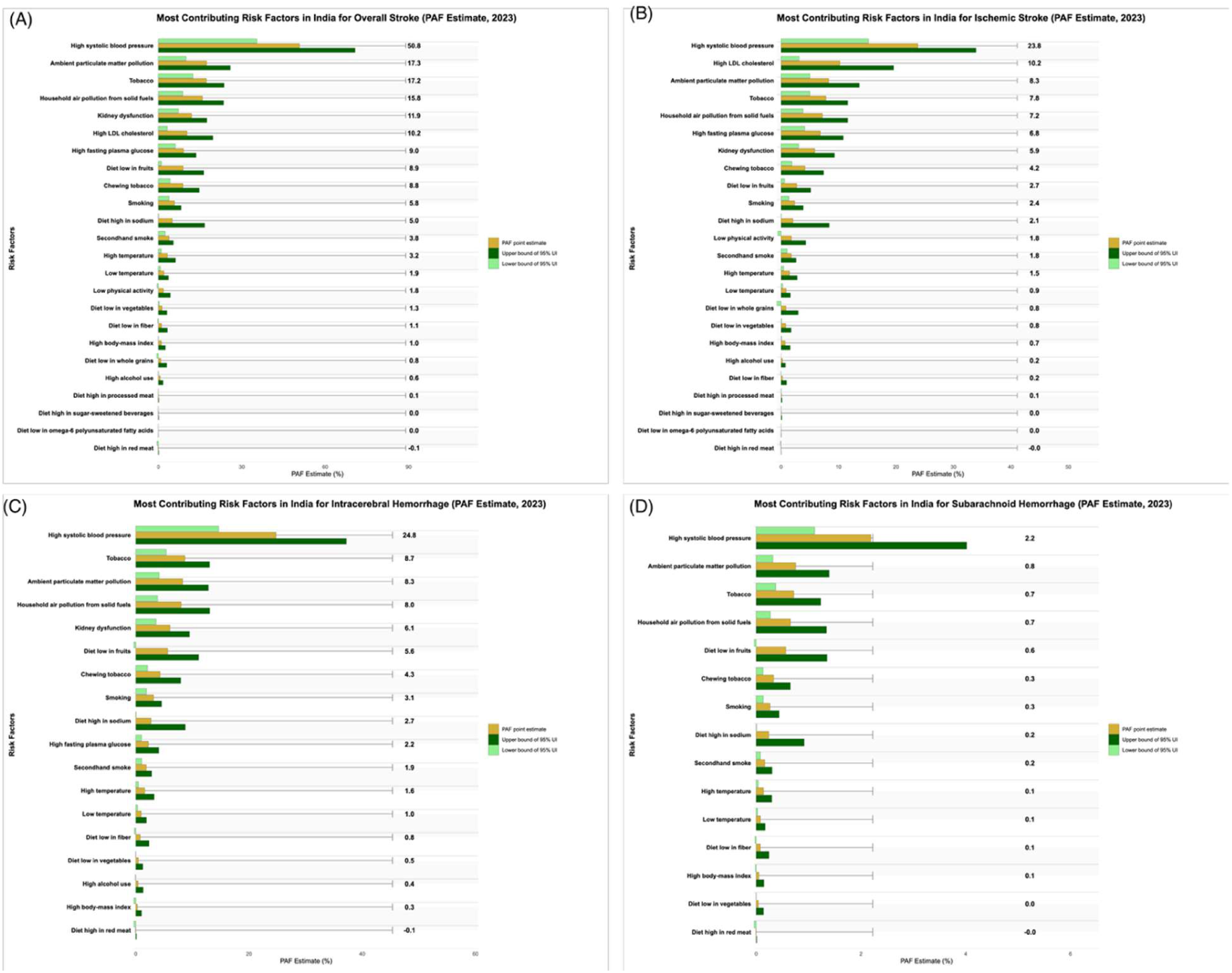
Most individually significant risk factors for total stroke (A), Ischaemic stroke (B), Intracerebral haemorrhage (C), and subarachnoid hemorrhage (D), as measured by the PAF of stroke DALYs attributable to the risk factors, for both sexes; DALYs=disability-adjusted life-years. PAF=population attributable fraction

**Figure 6.**
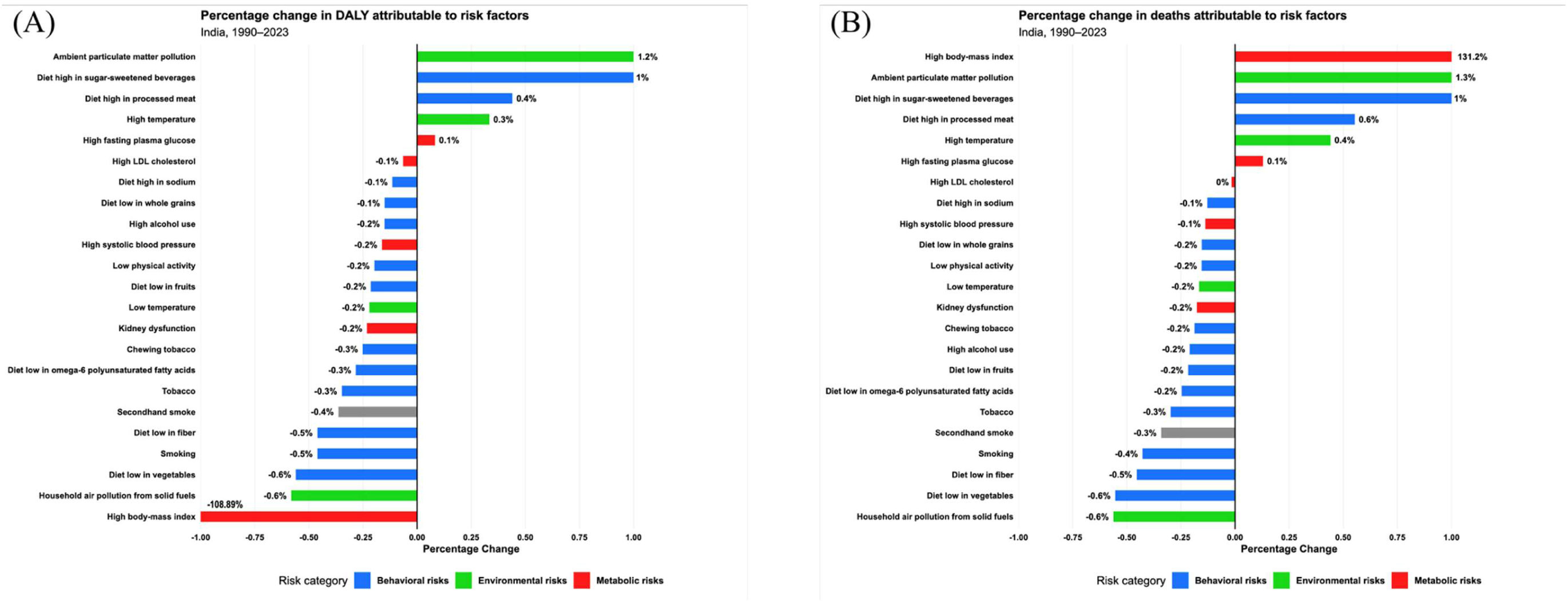
(A-B): (A)Trends in the PAF of stroke Deaths due to risk factors, for both sexes, 1990–2023; (B) Trends in the PAF of stroke DALYs due to risk factors, for both sexes, 1990–2023. Data in parentheses are 95% uncertainty intervals. DALYs=disability-adjusted life-years. PAF=population attributable fraction

### Projections of Stroke Burden to 2035

Based on current trends, the burden of stroke in India is expected to continue increasing over the coming decade. By 2035, the age-standardized incidence rate is projected to reach approximately 134.53 per 100,000 (95% CI: 122.10–150.30), while the mortality rate may rise to 88.99 per 100,000 (95% CI: 82.02–94.52). Similarly, the age-standardized DALY rate is projected to increase to 1,911.03 per 100,000 (95% CI: 1,758.18–2,022.15).

These projections suggest that, without strengthened prevention strategies and improved control of key risk factors, the overall burden of stroke in India is likely to remain substantial and may continue to rise in the future (**Fig. 7**).

**Figure 7:**
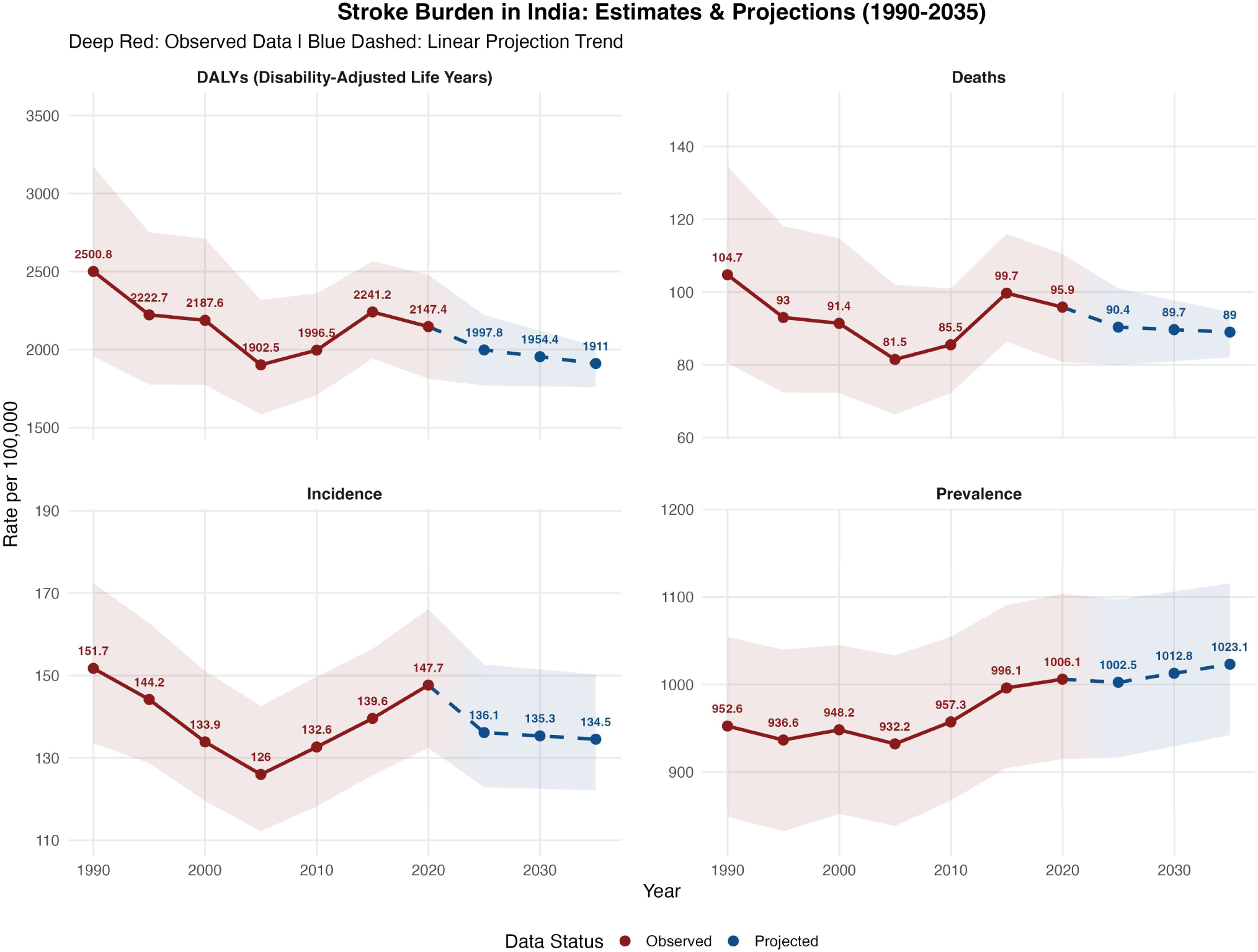
Trends and projections of stroke burden in India based on Global Burden of Disease (GBD) estimates (1990–2035)

## Discussion

This study provides a comprehensive assessment of the evolving burden of stroke in India from 1990 to 2023, demonstrating a substantial rise in the absolute number of incident cases, prevalent cases, deaths, and DALYs. Alongside long-term temporal trends, the analysis highlights important state-level variations in stroke burden and associated risk factors, and presents projections up to 2035. Together, these findings offer critical insights into the changing epidemiological landscape of stroke in India and underscore its growing public health importance.

The observed increase in stroke burden aligns with global patterns.^42–44^ Although many regions have seen improvements in prevention and clinical care, the overall number of stroke events has continued to rise worldwide.^45,46^ This trend is largely driven by population growth, demographic aging, and increasing exposure to metabolic and behavioral risk factors. In countries like India, which are undergoing rapid epidemiological transition, non-communicable diseases have become the leading contributors to morbidity and mortality, with stroke emerging as a major component of this burden. ^47^

A key finding of this study is the marked geographic variation in stroke burden across Indian states. These differences likely reflect variations in socioeconomic development, healthcare infrastructure, environmental exposures, and the distribution of modifiable risk factors. Such heterogeneity underscores the importance of subnational analyses in large, diverse countries like India. Identifying high-burden states is essential for prioritizing interventions and strengthening healthcare systems, particularly in resource-limited settings.

Modifiable risk factors play a central role in shaping the burden of stroke. Hypertension, diabetes, tobacco use, dietary patterns, and air pollution remain major contributors. Rapid urbanization and lifestyle changes have further increased the prevalence of these risk factors. Effective control of these determinants is therefore critical for reducing stroke incidence and improving population health outcomes. Public health strategies targeting cardiovascular risk factors have significant potential to curb the rising burden of stroke in India.

The inclusion of projections up to 2035 adds an important dimension to this study. The findings suggest that, if current trends persist, the burden of stroke in India will continue to increase over the coming decade. These projections emphasize the urgency of implementing effective prevention strategies and strengthening healthcare infrastructure to meet future demands.

### Strengths and Limitations

This study has several strengths. It provides a long-term analysis spanning more than three decades using standardized estimates from the Global Burden of Disease framework, which integrates multiple data sources to generate comparable estimates across time and regions. The availability of state-level estimates allows a detailed assessment of regional disparities, which is particularly important in a diverse country like India. Additionally, evaluating modifiable risk factors and incorporating future projections enhances the relevance of the findings for public health planning and policy development.

However, certain limitations should be acknowledged. GBD estimates rely on statistical modeling, particularly in regions with limited primary data, which may introduce uncertainty. Variations in data quality, diagnostic practices, and reporting systems across states may also affect the precision of subnational estimates. Furthermore, the use of secondary data limits the ability to examine individual-level clinical details such as treatment patterns and stroke severity. Finally, projections are based on current trends and assumptions, and future changes in healthcare policies or risk factor control may alter these trajectories.

### Public Health and Policy Implications

The findings have important implications for public health and policy in India. The rising absolute burden of stroke highlights the urgent need to strengthen primary prevention strategies, particularly targeting hypertension, diabetes, unhealthy diet, tobacco use, and air pollution. Expanding population-based screening programs and integrating them into primary healthcare systems will be crucial. Strengthening national initiatives such as the National Programme for Prevention and Control of Cancer, Diabetes, Cardiovascular Diseases, and Stroke (NPCDCS) can play a key role in reducing future burden^48^.

Improving public awareness of stroke symptoms and risk factors is equally important for early recognition and timely treatment.^49^ At the same time, strengthening acute stroke care infrastructure—including access to stroke units, emergency response systems, and advanced therapies such as thrombolysis and thrombectomy—can significantly reduce mortality and disability.^50^ Given the observed regional disparities, policies should be tailored to state-specific needs, with greater focus on high-burden areas. Investments in workforce training, surveillance systems, and rehabilitation services will be essential.

## Conclusion

In conclusion, this study highlights the growing burden of stroke in India over the past three decades, despite modest improvements in age-standardized rates. Significant regional disparities persist, driven by differences in socioeconomic conditions, healthcare access, and exposure to modifiable risk factors. Projections indicate that the burden is likely to continue increasing through 2035 if current trends persist. Addressing this challenge will require coordinated efforts focused on prevention, early detection, improved acute care, and expanded rehabilitation services. Region-specific strategies will be essential to reduce the health, social, and economic impact of stroke in India.

## Author contributions

MN, PT & BA: Study screening, Data curation & analysis, UJ & AJ: extraction and validation of data, KKP & ADS: Formal analysis, AKP & DV: writing, reviewing & editing. PK: Conceptualization, Formal analysis, Writing – original draft

## Data availability statement

The original contributions presented in the study are included in the article/Supplementary material; further inquiries can be directed to the corresponding author.

## Ethics statement

This study used publicly available, de-identified secondary data from the GBD Study. As no individual-level identifiable data were involved, ethical approval and informed consent were not required.

## Acknowledgement

None

